# Effects of Telehealth-based Lifestyle Modification on Cardiorespiratory Fitness Among Individuals with High-normal or High Blood Pressure: Randomized Controlled Trial

**DOI:** 10.64898/2025.12.29.25342521

**Authors:** Hongyi Du, Xuezhu Sun, Shumin Zhang, Baolu Wang, Meice Tian, Juan Deng, Liu Du, Peng Wang, Mingzhao Du, Xu Yang, Yutian Zhu, Qi Xie, Zijing Zeng, Xue Feng

**Author notes:** **Correspondence to:** Xue Feng, MD, Center for Lifestyle Medicine, Fuwai Hospital, National Center for Cardiovascular Diseases, Chinese Academy of Medical Sciences and Peking Union Medical College, 167A Beilishi Rd, Xi Cheng District, Beijing 100037, China.

## Abstract

**BACKGROUND:** Cardiorespiratory fitness (CRF) is a strong predictor of cardiovascular risk, yet scalable approaches to improve CRF among adults with high-normal or high blood pressure (BP) remain limited. Telehealth lifestyle programs may address this gap, but trials with rigorous CRF endpoints are scarce.

**METHODS:** We conducted a two-center, assessor-blinded, randomized controlled trial in Beijing and Shenzhen, China, from July 2022 to September 2024. Adults with high-normal or high BP were enrolled and randomized 1:1 to a telehealth-based lifestyle modification program (Tele-LM) or enhanced usual care (EUC), stratified by BP category. Tele-LM combined app-based dietary coaching, individualized aerobic and resistance prescriptions, home BP monitoring, and motivational interviewing. EUC comprised standardized lifestyle counseling, home BP monitoring, and educational resources. The primary outcome was change in peak oxygen uptake (VO_2peak_) at 3 months. Secondary outcomes included changes in VO_2peak_ at 9 months, home, ambulatory and office BP, lifestyle behaviors, metabolic parameters, and medication use. Analyses followed the intention-to-treat principle with multiple imputation for missing data.

**RESULTS:** Among 424 randomized participants (mean age 50 years; 65% men), 314 completed the 3-month assessment and 377 completed the 9-month assessment. At 3 months, the adjusted between-group difference in VO_2peak_ change favored Tele-LM by 1.5 mL/kg/min (95% confidence interval [CI], 0.6 to 2.3; *P*=0.001); at 9 months, a smaller difference persisted (1.0 mL/kg/min [95%CI, 0.1 to 1.9]; *P*=0.027). Subgroup analyses by BP category (randomization strata) revealed no significant heterogeneity for VO_2peak_. Tele-LM produced greater 3-month reductions in home BP (systolic BP −3.3 mmHg [95% CI, −5.3 to −1.4]; diastolic BP −2.6 mmHg [95%CI, −3.9 to −1.3]) and in 24-hour ambulatory BP (systolic BP −3.3 mmHg [95%CI, −6.0 to −0.6]; diastolic BP −1.9 mmHg [95%CI, −3.6 to −0.3]); between-group differences generally attenuated by 9 months. Adverse events were infrequent and similar between groups; no serious adverse events were observed. Average antihypertensive medication intensity was modestly lower in the Tele-LM (−0.27 agents [95% CI, −0.44 to −0.10]). Findings were consistent across prespecified sensitivity analyses.

**CONCLUSIONS:** In adults with high-normal or high BP, telehealth lifestyle modification with minimal in-person contact significantly improved CRF and modestly lowered BP, supporting its scalability as a complement to routine hypertension care.

**REGISTRATION:** URL: https://www.clinicaltrials.gov; Unique identifier: NCT05528068.

**Clinical Perspective:** *What Is New?:*

- This randomized trial evaluated a comprehensive telehealth-based lifestyle modification program, combining app-based dietary coaching, individualized aerobic and resistance prescriptions, wearable integration and motivational interviewing, in adults with high-normal blood pressure or well-controlled hypertension.
- Telehealth-based lifestyle modification significantly improved cardiorespiratory fitness and modestly reduced blood pressure compared with enhanced usual care.

*What Are the Clinical Implications?:*

- Telehealth lifestyle programs can extend preventive care to large at-risk populations with minimal in-person contact.
- By reducing reliance on pharmacologic intensification and conserving healthcare resources, telehealth-based strategies offer a cost-effective model for scalable cardiovascular prevention.
- Sustaining adherence remains essential to maximize long-term benefits.

## Introduction

Hypertension and high-normal blood pressure (BP) are highly prevalent and contribute substantially to cardiovascular morbidity and mortality. In China alone, hundreds of millions live with elevated BP.^1^ Despite guideline-recommended lifestyle measures—sodium restriction, a plant-rich diet, regular physical activity, weight control and adherence to antihypertensive therapy,^2,3^ engagement remains suboptimal in routine care. The COVID-19 pandemic further disrupted access to in-person counseling, underscoring the need for scalable, remotely delivered interventions.^4^

Cardiorespiratory fitness (CRF) is a powerful, independent predictor across the BP risk spectrum.^5,6^ Center-based exercise programs can improve CRF and lower BP but are constrained by travel, time, and resource demands.^7,8^ Although the dose–response relationship between physical activity and fitness is well established,^9^ practical tools to help large numbers of patients implement and sustain lifestyle changes outside clinic settings remain limited.

Telehealth and wearable-enabled lifestyle programs provide remote dietary and exercise support, objective activity tracking, and home BP monitoring with minimal in-person contact.^10–12^ Several trials have suggested modest improvements in fitness, BP or glycemic control.^13–16^ However, results have been inconsistent and durability beyond short intervention periods is unclear. Notably, whether telehealth lifestyle programs can improve CRF and confer additional cardiovascular benefits in adults with high-normal or well-controlled hypertension remains uncertain.

To address this gap, we conducted a randomized controlled trial to evaluate whether a low-touch telehealth-based lifestyle modification program (Tele-LM) improves CRF versus enhanced usual care (EUC) among adults with high-normal or high BP. We hypothesized that the telehealth-based approach will elicit clinically meaningful improvements in CRF, augment BP control and promote healthier lifestyle behaviors. The primary objective was to compare 3-month changes in peak oxygen uptake (VO_2peak_) measured by cardiopulmonary exercise testing (CPET).

## Methods

### Trial Design and Participants

This was a two-center, assessor-blinded, stratified, randomized controlled trial in Beijing and Shenzhen, China. The study was approved by the institutional review board at Fuwai Hospital (Beijing, China) and Fuwai Shenzhen Hospital (Shenzhen, China).

Adults aged 18–70 years with untreated high-normal BP (office systolic blood pressure [SBP] 120–139 mmHg and/or diastolic blood pressure [DBP] 80–89 mmHg) or high BP (office BP ≥140/90 mmHg or current antihypertensive therapy)^17^ were enrolled from July 2022 through December 2023, and then followed until September 2024. Patients were excluded for any of the following: acute myocardial infarction; tachyarrhythmia; pulmonary oedema; severe aortic stenosis; other serious cardiac or respiratory disorders; being in the acute phase of a cardiovascular or cerebrovascular event; poorly controlled hypertension (>160/100 mmHg despite treatment); physical limitations precluding CPET (e.g., fractures or unstable joints); skin disease at wearable-device placement site; active psychiatric illness or epilepsy impairing voluntary movement; permanent pacemaker implantation; and pregnancy or intent to become pregnant. All participants provided written informed consent.

### Randomization and Masking

Eligible participants were randomly assigned in a 1:1 ratio to either the Tele-LM or the EUC using a computer-generated randomization sequence created in R software, stratified by BP category (high-normal and high). The randomization list was prepared by an independent statistician not involved in participant enrollment or intervention delivery. Group assignments were concealed using sequentially numbered, opaque, sealed envelopes until the time of allocation. Outcome assessors and statisticians were blinded to group assignments, whereas participants and intervention staff were not blinded due to the nature of the behavioral intervention.

### Interventions

#### Tele-LM

Participants received a low-touch, multi-component telehealth lifestyle program delivered through an interactive smartphone application (Healthy Lifestyle App, OPPO Mobile Telecommunications Co., Guangdong, China). The program included individualized lifestyle prescriptions, dietary and exercise tracking reminders, education, home BP monitoring with automated feedback, and brief remote coaching. Content was aligned with guideline-recommended non-pharmacological management of hypertension, and recommendations were tailored to baseline characteristics and participant preferences.

This digital health platform was developed collaboratively by OPPO Co. under expert supervision from Fuwai Hospital, Beijing, China, comprising a mobile app for participants and a web-based interface for healthcare providers.

##### Dietary coaching

Dietary prescriptions were automatically generated based on participants’ semi-quantitative food frequency questionnaire (FFQ) responses and Dietary Approaches to Stop Hypertension (DASH) principles,^18,19^ including caloric intake, dietary structure, preferred foods, and recommendations for reduced salt and alcohol consumption. Prescriptions were reviewed and adjusted by dietitians. During the intervention, participants photographed meals, and dietitians provided weekly feedback emphasizing increased consumption of fruits, vegetables, and whole grains, limitation intake of sodium and saturated fats, and adherence to DASH dietary pattern.

##### Supervised exercise

Individualized aerobic and resistance exercise plans were automatically generated based on baseline CPET results, combining specific aerobic and resistance exercise modalities. Participants were instructed to complete at least 150 minutes per week of moderate-intensity aerobic exercises and two sessions of resistance training weekly. Resistance training sessions could be completed by following instructional videos provided within the app. Wearable devices recorded heart rate and step counts, and adherence was logged through the application. Participants received exercise reminders, and recorded exercise duration, type and fatigue level. Automated alerts were generated if heart rate exceeded predetermined safety thresholds. Physical therapists provided online supervision, guidance, and weekly feedback based on interactive user engagement, adjusting exercise plans accordingly.

##### BP monitoring and feedback

Participants were provided with validated home BP monitors and instructed to perform home BP monitoring at least three days per week, following standardized procedures. Readings were synced to the app and reviewed remotely. Automated alerts were triggered when a home BP value was markedly elevated (>180/110 mmHg) or abnormally low (SBP <100 mmHg), prompting participants to seek medical attention within three days.

##### Motivational interviewing

Motivational interviewing sessions via phone calls or videoconferencing were conducted if adherence to dietary or exercise logging fell below 50%, aiming to reinforce behavioral changes. Clinical psychologists provided counseling to improve eating behaviors, increase physical activity, address individual barriers, and support long-term behavior change.

Participants in the Tele-LM actively engaged in these interventions for three months, followed by a self-managed period of an additional six months (up to nine months), with data on self-monitoring collected through the app.

#### EUC

Participants received standardized counseling at enrollment from dietitian on DASH diet and from physical therapist on guideline-based physical activity. They were provided a validated home BP monitor and a wearable device to record BP and step counts. Automated safety alerts were triggered for predefined out-of-range BP values, instructing participants to seek medical advice from their usual clinicians. The EUC did not include individualized dietary and exercise prescriptions, tailored feedback, or remote coaching. Antihypertensive therapy was managed by participants’ usual clinicians per guideline-based care in both groups. The protocol neither mandated nor restricted medication initiation, titration, or deprescription.

### Measurements and Assessments

#### Cardiorespiratory fitness

CPET was performed to assess CRF using the same standard operating procedures at both study sites, employing an MasterScreen® cycle ergometer (Vyaire Medical, Hoechberg, Germany), in accordance with international guidelines.^20^ Each test began with a 3-minute warm-up at five watts, followed by an individualized ramp protocol targeting VO_2peak_ within 8–12 minutes. Quality criteria included exceeding the first ventilatory threshold and achieving a respiratory exchange ratio (RER) >1.1. Heart rate was continuously recorded via electrocardiographic monitoring. CPETs were conducted at baseline, 3 months, and 9 months.

#### Blood pressure

Office BP was measured at each visit. Two readings were taken per visit, and the average was recorded if the difference was <10 mmHg. If the difference exceeded 10 mmHg, additional readings were obtained until two stable values were achieved. Home BP monitoring was conducted at least 3 days per week, with measurements taken twice daily (morning and evening), using a validated device (U36T; Omron Healthcare Co., China). Ambulatory BP monitoring (ABPM) was performed at baseline, 3 months, and 9 months using a validated device (M2; Raycome Health Technology Co., ShenZhen, China). The monitor was programmed to record BP hourly during waking hours and every two hours during sleep. At least 70% of scheduled readings were required for an ABPM session to be considered valid. Mean 24-hour BP was calculated as the average of all successful readings. Both ABPM and home BP monitors were provided by the study team and connected via Bluetooth to the participant’s smartphone app, which automatically transferred readings to the study database.

#### Anthropometric and metabolic parameters

Body weight and height were assessed at each visit to compute body mass index (BMI). Body composition, specifically fat mass as a percentage of total body weight, was measured by bioelectrical impedance analysis (InBody 770; InBody Co., Korea). Fasting venous blood was collected at baseline, 3 months, and 9 months to measure total cholesterol, low-density lipoprotein cholesterol (LDL-C), high-density lipoprotein cholesterol (HDL-C), triglycerides, and fasting glucose. Tests were performed in local certified laboratories.

#### Dietary assessment

Dietary intake was assessed using a semi-quantitative FFQ and evaluated one-on-one by trained dietitians at each study visit. A modified DASH score was calculated based on a 10-component scoring system adapted from Folsom et al.,^21^ covering grains; fruits; vegetables; nuts, seeds, and legumes; dairy; meat; fat; unsaturated fat; sweets; and sodium. Each component was scored 0, 0.5, or 1 based on adherence to DASH dietary recommendations (Table S1), with a total score ranging from 0 to 10, where higher scores indicated better adherence.

#### Physical activity

Physical activity was assessed online at each study visit using the International Physical Activity Questionnaire–Short Form (IPAQ-SF).^22^ Data were processed according to the IPAQ scoring protocol. Weekly activity levels were expressed as metabolic equivalent-minutes per week (MET-min/ week), calculated as duration × frequency × assigned MET intensity. Moderate to vigorous physical activity (MVPA) was derived as the sum of MET-minutes/ week from reported moderate- and vigorous-intensity activities, and walking activity was quantified independently according to the IPAQ-SF scoring protocol.

#### Objective activity monitoring

Participants wore a wrist-worn device (OWW211 OPPO Watch 3 Pro; OPPO Mobile Telecommunications Co., Guangdong, China) to capture steps continuously during the study. Because pre-enrollment data were unavailable, mean daily step counts were summarized for months 0–3 and months 4–9 as absolute levels and analyzed as a complement to questionnaire-based measures.

#### Medication use

Medication use was assessed at baseline and follow-up visits using structured online questionnaires. Information was collected on the number and classes of antihypertensive agents prescribed. All medication changes were recorded, and medication intensity was summarized as the number of agents.

#### Adherence

Adherence was evaluated over two assessment windows (months 0–3 and 4–9). Participants were classified as adherent if they met the predefined weekly target in at least 60% of weeks within each window. The exercise target was defined as either completing ≥150 minutes of aerobic activity per week or fulfilling the full prescription of ≥150 minutes of aerobic activity plus two resistance sessions per week. Dietary adherence was defined as submitting dietary records on at least two days per week through the app. Home BP monitoring adherence was defined as measuring BP on at least three days per week, with values uploaded via the app.

#### Adverse events

Adverse events (AEs) were monitored through monthly telephone follow-ups in both groups. An adverse event was defined as any unfavorable medical occurrence during the trial.

Additional exploratory measurements prespecified in the protocol were collected but are not included in this report. Detailed assessment procedures are provided in Supplemental Methods (Protocol).

### Outcomes

The primary outcome was the change in CPET-measured VO_2peak_ from baseline to 3 months. Secondary outcomes included changes in VO_2peak_ at 9 months and other CPET measures (maximal heart rate [HRmax], peak workload, and RER) from baseline to 3 and 9 months; changes in all BP variables, including home BP (mean morning, mean evening, and overall SBP/DBP), ambulatory BP (24-hour, daytime, nighttime SBP/DBP), and office SBP/DBP; changes in BMI and fat mass percentage; changes in metabolic parameters (fasting glucose, total cholesterol, LDL-C, HDL-C, and triglycerides); changes in DASH score, IPAQ-SF–derived MVPA and walking activity; objective activity as daily step count during months 0–3 and 4–9; medication use (number and pattern of antihypertensive agents, categorized as unchanged/reduced vs intensified by study end); adherence (prespecified metrics for exercise, dietary logging, and home BP monitoring); and number of adverse events. Unless otherwise specified, secondary outcomes were analyzed as change from baseline to 3 months and 9 months.

### Impact of COVID-19 on Study Procedures

In December 2022, new enrollments and in-person data collection were halted due to the COVID-19 pandemic. This led to subsequent adjustments to the study protocol to maximize data collection from enrolled participants. If the 3-month CPET could not be completed due to COVID-related disruptions, the 6-month CPET was obtained per a prespecified contingency and used as a substitute for the 3-month endpoint in the primary analysis. These changes were implemented consistently across study centers and documented to ensure accurate interpretation of results.

### Sample Size

Sample size estimation was based on previous trials,^23,24^ with an expected between-group difference in VO_2peak_ change of 2.0 mL/kg/min among participants with high BP and 3.0 mL/kg/min among those with untreated high-normal BP, assuming a common standard deviation of 4.5 mL/kg/min. Because the primary endpoint was evaluated separately within two stratification groups (high-normal and high BP), a Bonferroni-adjusted two-sided alpha level of 0.025 was applied to control type I error. With 90% power, the required sample size was calculated to be 176 participants per group (52 with high-normal BP and 124 with high BP). Allowing for an anticipated dropout rate of 20%, the recruitment target was set at 212 participants per group, yielding a total sample size of 424.

### Statistical Analysis

Main analyses followed the intention-to-treat (ITT) principle. Continuous variables were summarized as mean ± standard deviation (SD), and categorical variables as counts and percentages. The primary analysis compared between-group changes in VO_2peak_ from baseline to 3 months using general linear models, adjusting for baseline VO_2peak_, BP category, study center, age, sex, education level, per-capita household income, and smoking status. Unadjusted and adjusted mean differences with 95% confidence intervals (CIs) and *P*-values were reported. Secondary outcomes were analyzed similarly, with adjustment for baseline values of the dependent variable. To provide a scale-independent measure of effect size, standardized mean differences (SMDs) were calculated for all continuous outcomes, defined as the adjusted mean difference divided by the pooled standard deviation. Prespecified subgroup analyses compared intervention effects across strata defined by BP category (randomization strata), age, sex, center, and smoking status, adjusting only for baseline values of the dependent variable. Interaction between intervention assignment and each subgroup variable was assessed by adding a multiplicative interaction term to the general linear models. Sensitivity analyses were then conducted to assess the robustness of findings. These included complete-case (CC) analyses and a per-protocol (PP) population restricted to participants with high adherence. For the PP analysis, Tele-LM participants were excluded if they achieved the planned weekly exercise duration in fewer than 50% of weeks during the first 3 months. If the 3-month outcome was missing, the corresponding 6-month value was used as a substitute when available. Remaining missing data were imputed using multiple imputation by chained equations (100 imputations), under the assumption that data were missing at random conditional on observed covariates and outcomes.^25^ The imputation model included all outcome and stratification variables. All analyses were conducted using R version 4.4.2 (R Foundation for Statistical Computing, Vienna, Austria). Figures were generated with the ggplot2 package. Two-tailed *P*-values <0.05 were considered statistically significant.

## Results

### Participant Flow and Baseline Characteristics

Participants recruitment started on July 2022 and the last one completed the study on September 2024. Among 466 individuals screened, 424 were enrolled and randomly assigned to EUC or Tele-LM (212 per group), after stratification by BP category (high-normal or high). Follow-up visits at 3 months were completed by 160/212 (75.5%) in EUC and 154/212 (72.6%) in Tele-LM; at 9 months by 193/212 (91.0%) and 184/212 (86.8%), respectively. To address missed visits due to COVID-19, a prespecified contingency allowed substitution of 6-month CPET values for 3-month values when visits were disrupted; these substitutions were included in the primary ITT analysis (EUC n=31; Tele-LM n=29; Figure 1).

**Figure 1.**
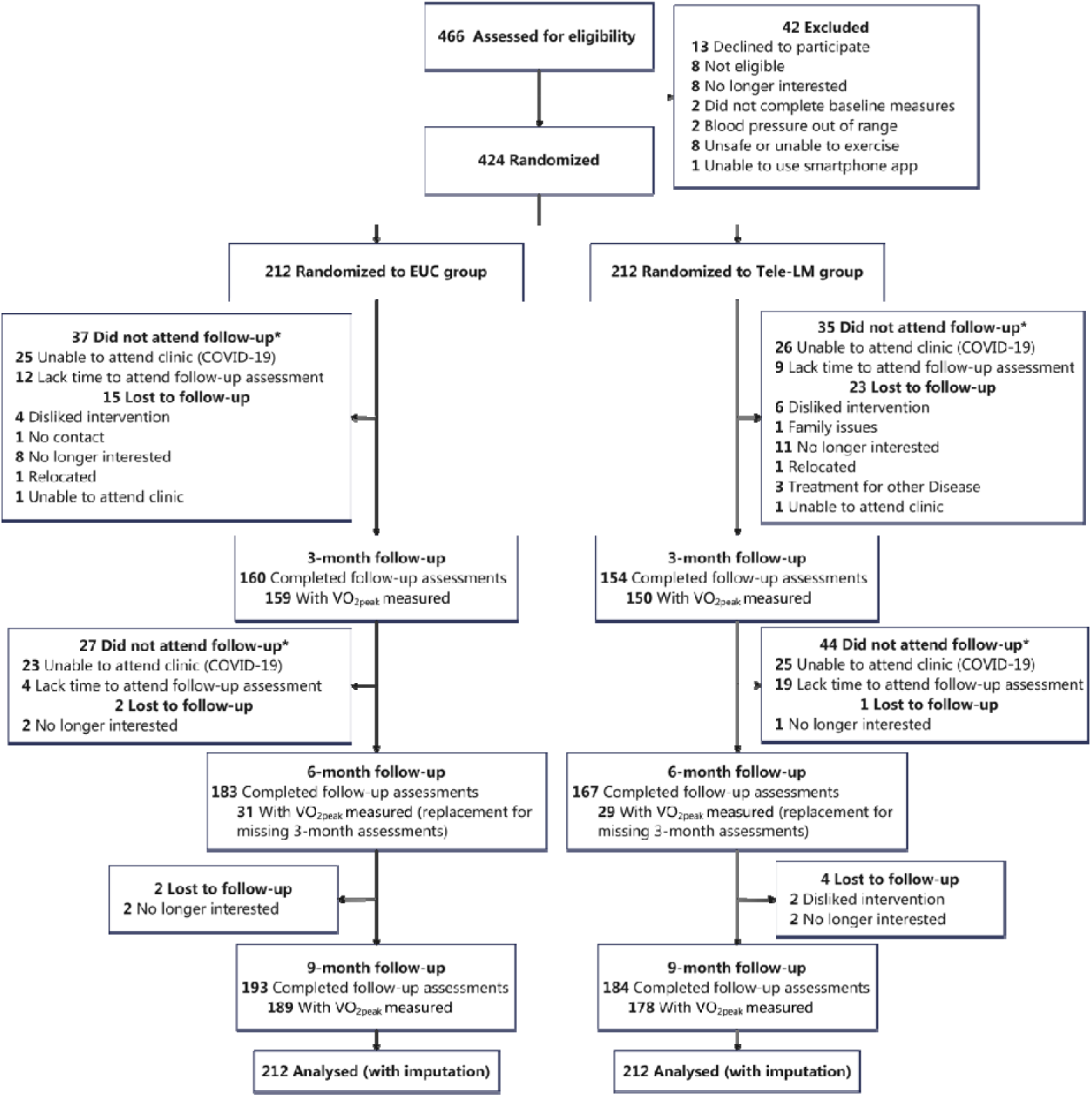
CONSORT Flow Diagram. Participants were assessed for eligibility, randomized, and followed through 3, 6, and 9 months. “Completed follow-up assessments” refers to complete study visits at each time point. Numbers with VO_2peak_ measured indicate those with available cardiopulmonary exercise testing (CPET) data prior to imputation. At 6 months, VO_2peak_ testing was performed in a subset as replacement for the missing of 3-month assessments. All randomized participants (N=424) were included in the intention-to-treat analysis using multiple imputation for missing data. EUC indicates enhanced usual care; Tele-LM, telehealth-based lifestyle modification program; VO_2peak_, peak oxygen uptake. *\** Participants who did not attend a given follow-up visit may have attended subsequent visits.

Baseline characteristics were well balanced between groups (Table 1). Participants averaged 50 years of age and 65.1% were men. Over half had family history of hypertension and 13.7% had diabetes. At baseline, office BP was slightly higher in Tele-LM, while home and ambulatory BP values were similar. Regarding antihypertensive medication use, about half were not taking antihypertensives at enrollment, and roughly one-third used a single agent. Use of more than two agents was infrequent.

**Table 1.** Baseline Demographic and Clinical Characteristic.

| **Characteristic** | **All**<br>**participants**<br>**(N=424)** | **EUC**<br>**(N=212)** | **Tele-LM**<br>**(N=212)** |
| --- | --- | --- | --- |
| Age, years | 50.5 (10.9) | 51.0 (10.7) | 49.9 (11.2) |
| Male, n (%) | 276 (65.1) | 134 (63.2) | 142 (67.0) |
| Blood pressure category, n (%) |  |  |  |
| High-normal blood pressure | 129 (30.4) | 64 (30.2) | 65 (30.7) |
| High blood pressure | 295 (69.6) | 148 (69.8) | 147 (69.3) |
| Education level, n (%) |  |  |  |
| Middle school and below | 25 (5.9) | 8 (3.8) | 17 (8.0) |
| High school | 47 (11.1) | 23 (10.8) | 24 (11.3) |
| College degree | 257 (60.0) | 127 (59.9) | 130 (61.3) |
| Graduate degree | 95 (22.4) | 54 (25.5) | 41 (19.3) |
| Per-capita household income monthly, yuan, n (%) |  |  |  |
| < 5000 | 62 (14.6) | 27 (12.7) | 35 (16.5) |
| 5000-10000 | 110 (25.9) | 59 (27.8) | 51 (24.1) |
| 10000-30000 | 177 (41.7) | 85 (40.1) | 92 (43.4) |
| > 30000 | 75 (17.7) | 41 (19.3) | 34 (16.0) |
| Smoking status, n (%) |  |  |  |
| Current | 82 (19.3) | 38 (17.9) | 44 (20.8) |
| Former | 44 (10.4) | 25 (11.8) | 19 (9.0) |
| Never | 298 (70.3) | 149 (70.3) | 149 (70.3) |
| Medical history, n (%) |  |  |  |
| Diabetes | 58 (13.7) | 32 (15.1) | 26 (12.3) |
| Family history of hypertension | 227 (53.7) | 111 (52.4) | 116 (55.0) |
| Coronary artery disease | 25 (5.9) | 13 (6.1) | 12 (5.7) |
| Stroke | 8 (1.9) | 3 (1.4) | 5 (2.4) |
| Chronic obstructive pulmonary disease | 2 (0.5) | 1 (0.5) | 1 (0.5) |
| Medication, n (%) |  |  |  |
| Diuretic | 27 (6.4) | 19 (9.0) | 8 (3.8) |
| β-Blockers | 86 (20.3) | 46 (21.7) | 40 (18.9) |
| ACE inhibitors/angiotensin receptor antagonists | 90 (21.2) | 46 (21.7) | 44 (20.8) |
| Calcium channel blockers | 54 (12.7) | 27 (12.7) | 27 (12.7) |
| Direct vasodilators | 7 (1.7) | 4 (1.9) | 3 (1.4) |
| Other antihypertensive drugs* | 11 (2.6) | 4 (1.9) | 7 (3.3) |
| Antiplatelet drug | 47 (11.1) | 27 (12.7) | 20 (9.4) |
| Hypoglycemic drug | 48 (11.3) | 27 (12.7) | 21 (9.9) |
| Lipid-lowering drug | 107 (25.2) | 62 (29.2) | 45 (21.2) |
| Antidepressant drug | 2 (0.5) | 2 (0.9) | 0 (0) |
| Anxiolytic drug | 3 (0.7) | 1 (0.5) | 2 (0.9) |
| Number of antihypertensive medications, n (%) |  |  |  |
| 0 | 230 (54.2) | 112 (52.8) | 118 (55.7) |
| 1 | 139 (32.8) | 72 (34.0) | 67 (31.6) |
| 2 | 44 (10.4) | 20 (9.4) | 24 (11.3) |
| 3 | 9 (2.1) | 6 (2.8) | 3 (1.4) |
| ≥4 | 2 (0.5) | 2 (0.9) | 0 (0.0) |
| **Anthropometrics** |  |  |  |
| BMI, kg/m<sup>2</sup> | 26.2 (3.9) | 26.3 (3.9) | 26.1 (3.8) |
| Fat mass, % | 28.3 (7.0) | 28.6 (7.0) | 28.2 (7.0) |
| **Cardiorespiratory Fitness** |  |  |  |
| VO<sub>2peak</sub>, mL/kg/min | 23.0 (5.8) | 22.9 (5.7) | 23.0 (5.9) |
| HRmax, bpm | 141.8 (21.6) | 139.9 (22.2) | 143.6 (21.0) |
| RER | 1.2 (0.1) | 1.2 (0.1) | 1.2 (0.1) |
| Peak workload, Watt | 138.6 (53.1) | 136.7 (57.9) | 140.8 (47.8) |
| **Blood Pressure, mmHg** |  |  |  |
| Home SBP | 128.6 (11.5) | 128.4 (12.4) | 128.9 (10.5) |
| Home DBP | 82.0 (7.0) | 81.7 (7.2) | 82.4 (6.8) |
| Morning home SBP | 128.1 (11.7) | 127.9 (12.3) | 128.5 (11.1) |
| Morning home DBP | 83.0 (7.4) | 82.5 (7.4) | 83.4 (7.4) |
| Evening home SBP | 128.9 (12.1) | 129.2 (12.7) | 128.6 (11.3) |
| Evening home DBP | 82.0 (7.9) | 81.8 (8.2) | 82.3 (7.3) |
| 24-hour ambulatory SBP | 129.2 (11.5) | 128.8 (12.3) | 29.6 (10.6) |
| 24-hour ambulatory DBP | 81.9 (7.0) | 81.2 (7.1) | 82.5 (6.8) |
| Daytime ambulatory SBP | 129.9 (12.0) | 130.0 (12.5) | 129.8 (11.5) |
| Daytime ambulatory DBP | 82.2 (6.9) | 81.7 (7.0) | 82.8 (6.8) |
| Nighttime ambulatory SBP | 126.4 (13.3) | 126.1 (14.4) | 126.7 (12.2) |
| Nighttime ambulatory DBP | 80.3 (8.2) | 79.8 (8.5) | 80.7 (7.8) |
| Office SBP | 131.6 (13.6) | 130 (13.7) | 133.3 (13.2) |
| Office DBP | 83.5 (10.3) | 82.2 (10.5) | 84.7 (10.1) |
| **Cardiometabolic Parameters** |  |  |  |
| Fasting glucose, mmol/L | 6.0 (1.3) | 5.9 (0.9) | 6.0 (1.6) |
| Total cholesterol, mmol/L | 4.7 (1.0) | 4.6 (1.1) | 4.8 (1.0) |
| LDL-C, mmol/L | 2.8 (0.8) | 2.8 (0.9) | 2.9 (0.8) |
| HDL-C, mmol/L | 1.4 (0.4) | 1.3 (0.3) | 1.4 (0.4) |
| Triglycerides, mmol/L | 1.6 (1.1) | 1.6 (1.0) | 1.6 (1.1) |
| **Lifestyle** |  |  |  |
| DASH score | 5.8 (1.8) | 5.9 (1.8) | 5.7 (1.8) |
| MVPA, MET-min/ week | 564.6 (1140.8) | 499.4 (1141.6) | 629.8 (1139.0) |
| Walk, MET-min/ week | 985.6 (933.2) | 979.8 (944.1) | 991.3 (924.3) |

Data are mean (SD) unless stated otherwise. EUC indicates enhanced usual care; Tele-LM, telehealth-based lifestyle modification program; ACE, angiotensin-converting enzyme; BMI, body mass index, calculated as weight in kilograms divided by height in meters squared; VO<sub>2peak</sub>, peak oxygen uptake; HR<sub>max</sub>, maximal heart rate; RER, respiratory exchange ratio; SBP, systolic blood pressure; DBP, diastolic blood pressure; LDL-C, low-density lipoprotein cholesterol; HDL-C, high-density lipoprotein cholesterol; DASH, dietary approaches to stop hypertension; MVPA, moderate to vigorous physical activity; MET, metabolic equivalent task.
\* Including Chinese herbal medicine, single-pill combination and other types of antihypertensive medication.

### Primary Outcome: Cardiorespiratory Fitness

At 3 months, VO_2peak_ (primary outcome) increased in the Tele-LM and remained essentially unchanged in the EUC (mean change +1.3 mL/kg/min [95% CI, 0.6 to 1.9] vs. −0.2 mL/kg/min [95% CI, −0.8 to 0.4]), yielding an adjusted between-group difference of 1.5 mL/kg/min (95% CI, 0.6 to 2.3; *P*=0.001). At 9 months, a smaller difference persisted (1.0 mL/kg/min [95% CI, 0.1 to 1.9], *P*=0.027) (Figure 2, Table S2). In prespecified analyses by BP category at 3 months, VO_2peak_ benefits were observed in both strata (high-normal BP 2.1 mL/kg/min [95% CI, 0.3 to 4.0], *P*=0.026; high BP 1.3 mL/kg/min [95% CI, 0.2 to 2.3], *P*=0.015); the interaction was not significant (*P*=0.393; Table S3). Secondary CPET metrics at 3 months also favored Tele-LM, including peak workload 12.6 W (95% CI, 6.0 to 19.2; *P*<0.001) and HRmax 6.7 bpm (95% CI, 3.1 to 10.3; *P*<0.001). RER values did not differ significantly (*P*=0.073), and prespecified CPET quality targets were consistently met, supporting adequate maximal effort and test validity in both groups (Table S2).

**Figure 2.**
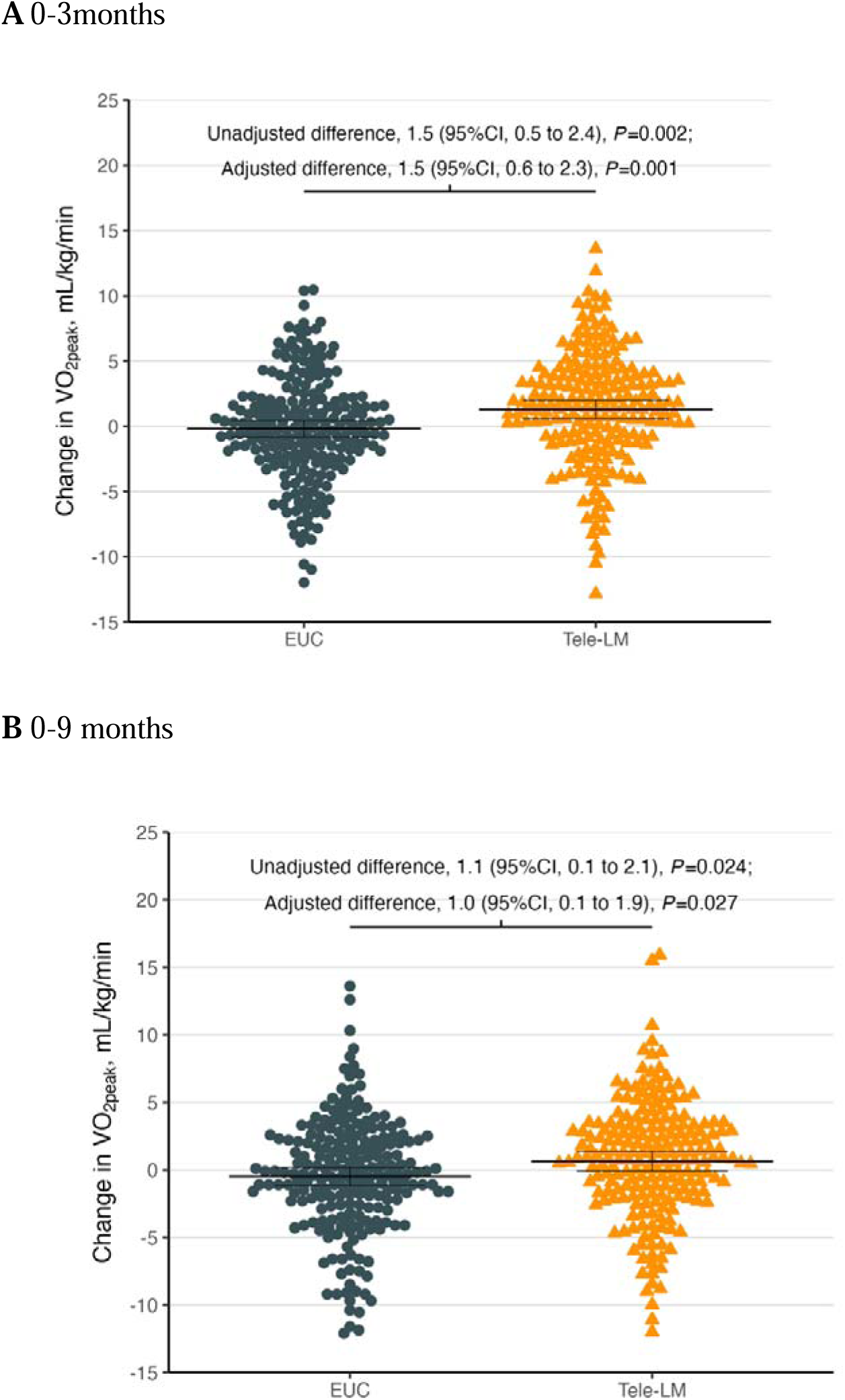
Change in VO_2peak_ at 3 (Primary Outcome) and 9 Months of Follow-up. Change in VO_2peak_ from baseline to 3- (**A**) and 9-months (**B**) follow-up is significantly greater in the Tele-LM compared with the EUC. Data are presented as mean (95% CI). The differences between groups with 95% CIs are displayed at the top. Adjusted differences reflect between-group changes in VO_2peak_ from baseline using general linear models, adjusting for baseline VO_2peak_, blood pressure category, study center, age, sex, education level, per-capita household income monthly, and smoking status. EUC indicates enhanced usual care; Tele-LM, telehealth-based lifestyle modification program; VO_2peak_, peak oxygen uptake.

### Blood Pressure and Cardiometabolic Outcomes

#### Blood pressure

At 3 months, Tele-LM achieved greater BP reductions than EUC. Adjusted between-group differences were −3.3 mmHg for home SBP (95% CI, −5.3 to −1.4; *P*<0.001) and −2.6 mmHg for home DBP (95% CI, −3.9 to −1.3; P<0.001). For 24-hour ambulatory BP, the adjusted differences favored Tele-LM (SBP −3.3 mmHg [95% CI, −6.0 to −0.6], *P*=0.019; DBP −1.9 mmHg [95% CI, −3.6 to −0.3], *P*=0.020). Office BP differences were smaller (SBP −1.9 mmHg [95% CI, −4.3 to 0.5], *P*=0.112; DBP −2.0 mmHg [95% CI, −3.7 to −0.4], *P*=0.018) (Figure 3). Patterns for morning/evening home BP and daytime/nighttime ambulatory BP were comparable. At 9 months, BP differences were generally attenuated (Table S2).

**Figure 3.**
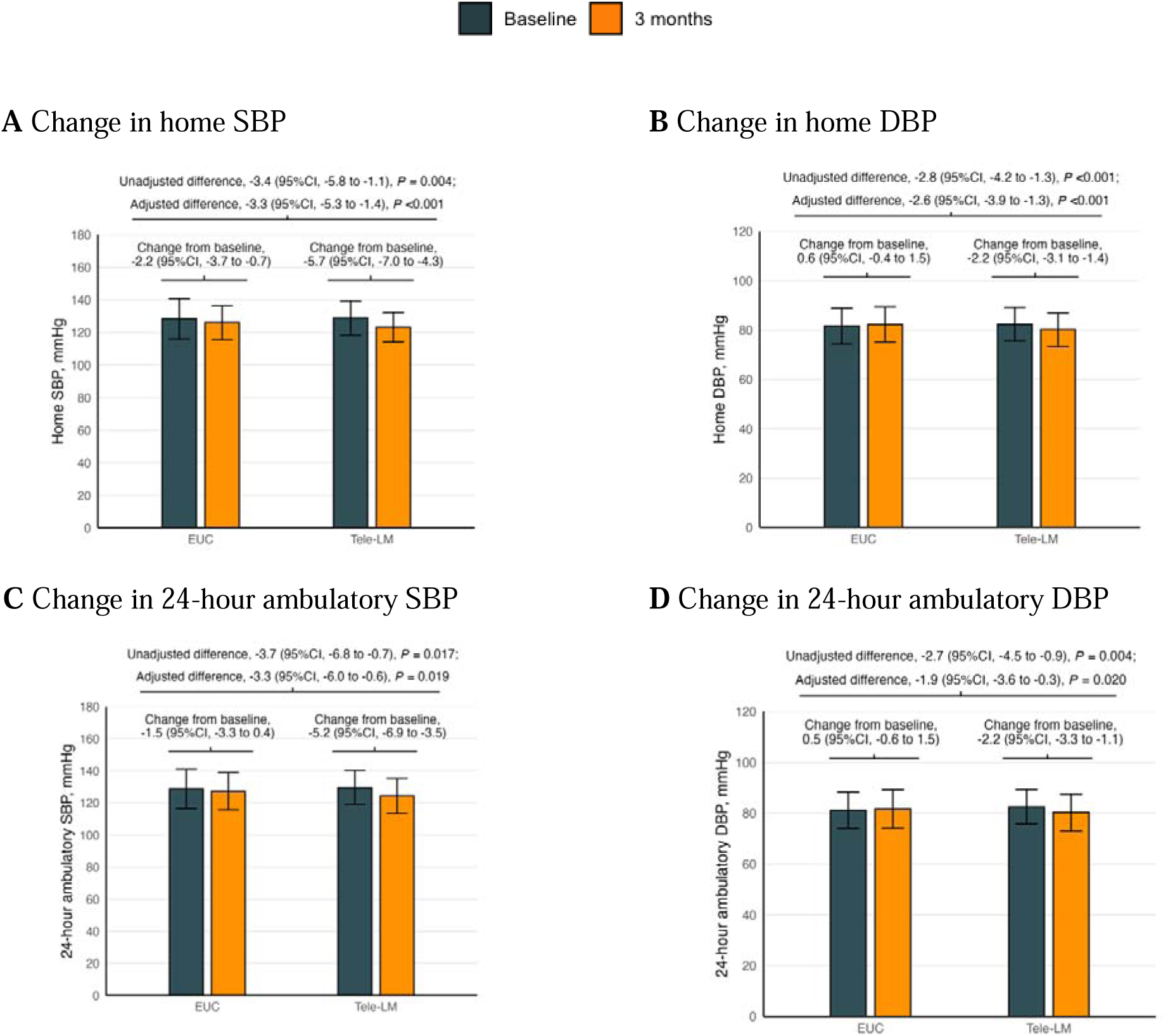
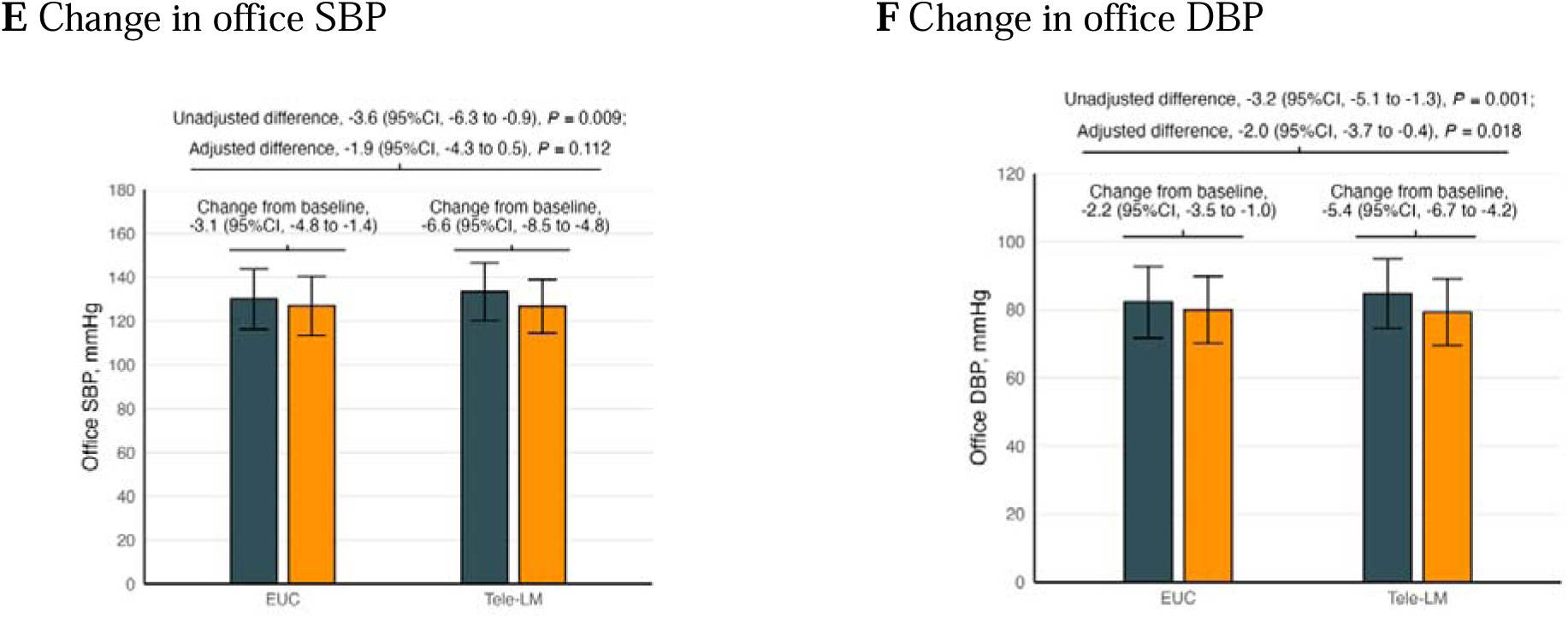
Changes in Home, Ambulatory and Office Blood Pressure at 3 Months of Follow-up. Adjusted differences reflect between-group differences in change from baseline in blood pressure using general linear models, adjusting for baseline blood pressure, blood pressure category, study center, age, sex, education level, per-capita household income monthly, and smoking status. Negative differences are in favor of the Tele-LM. EUC indicates enhanced usual care; Tele-LM, telehealth-based lifestyle modification program; SBP, systolic blood pressure; DBP, diastolic blood pressure.

#### Cardiometabolic and anthropometric outcomes

Changes in fasting glucose, triglycerides, LDL-C, HDL-C, and total cholesterol, BMI, and body fat percentage did not differ significantly between groups at 3 or 9 months (Table S2).

### Lifestyle Outcomes

At 3 months, diet quality improved more in Tele-LM, with an adjusted mean difference in DASH score of 0.6 points (95% CI, 0.3 to 0.9; *P*<0.001). MVPA also increased, with an adjusted mean difference of 324 MET-min/ week (95% CI, 65.9 to 582.8; *P*=0.014). At 9 months, improvements in both DASH score and MVPA were attenuated; however, the between-group difference in DASH score remained statistically significant, whereas the difference in MVPA was no longer significant. Walking activity, expressed as MET-min per week, did not differ significantly either between groups or within groups over time. Objective average daily steps from the wearable device showed no significant between-group differences for months 0–3 or months 4–9 (Table S2).

SMDs for all primary and secondary outcomes at 3 and 9 months were presented in Figure 4. At 3 months, effects were small to moderate across CRF, lifestyle, and BP outcomes, most of which had attenuated by 9 months. Metabolic biomarkers and body composition remained near null throughout follow-up.

**Figure 4.**
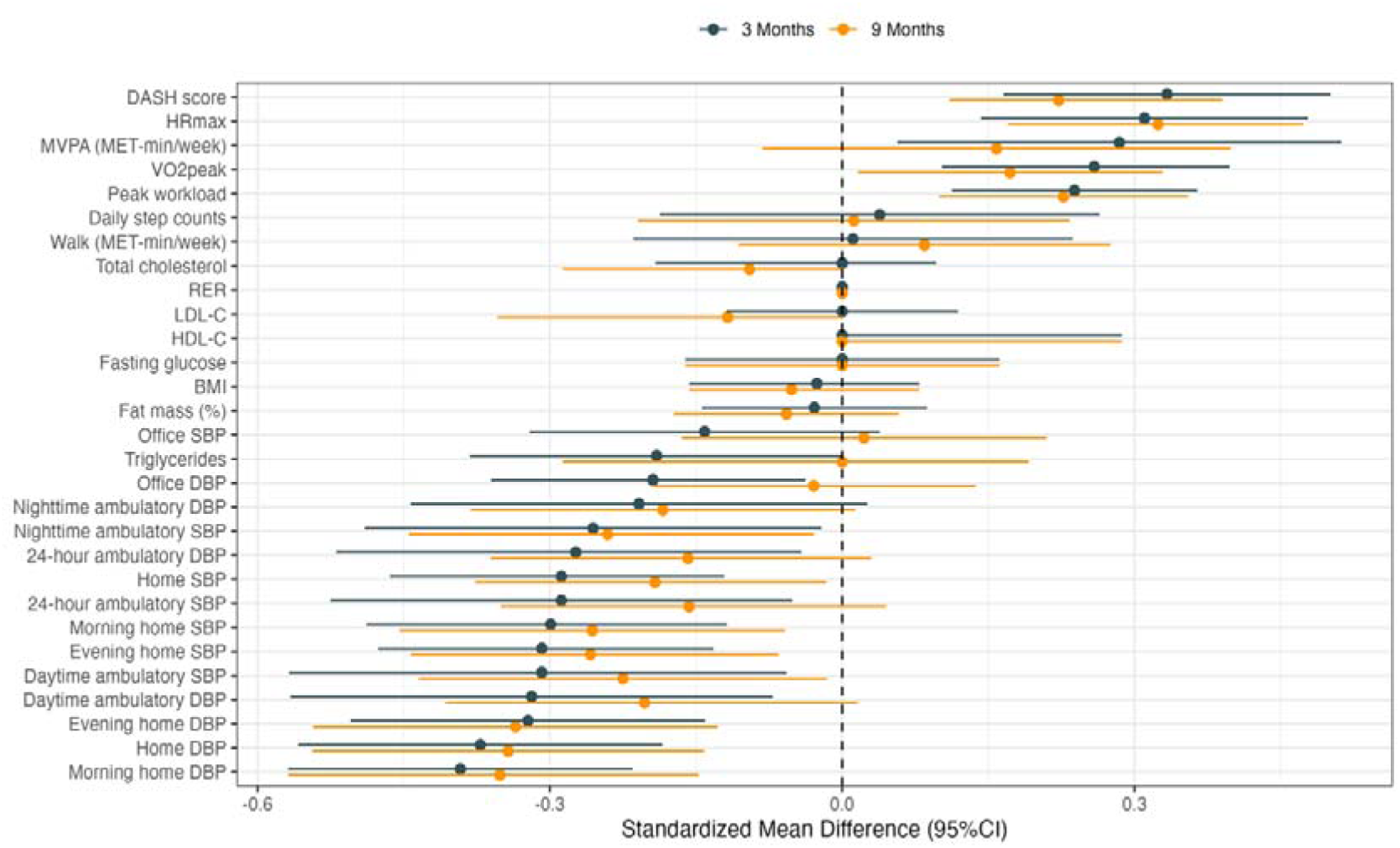
Standardized Mean Differences for Between-group Differences in Changes for Primary and Secondary Outcomes at 3 and 9 Months. Standardized mean differences (SMDs) were computed as the adjusted mean difference divided by the pooled standard deviation, providing a scalel-independent measure of effect size across variables. For higher-is-better outcomes (i.e., DASH score, HRmax, MVPA, VO_2peak_, Peak workload, Daily step counts, Walk, RER and HDL-C), positive SMD values indicate between-group difference in change favoring the telehealth-based lifestyle modification program (Tele-LM). For lower-is-better outcomes (i.e., BMI, Fat mass, Total cholesterol, LDL-C, Triglycerides, Fasting glucose, SBP and DBP), negative SMD values indicate between-group difference in change favoring the Tele-LM. DASH indicates dietary approaches to stop hypertension; HRmax, maximal heart rate; MVPA, moderate to vigorous physical activity; MET, metabolic equivalent task; VO_2peak_, peak oxygen uptake; RER, respiratory exchange ratio; LDL-C, low-density lipoprotein cholesterol; HDL-C, high-density lipoprotein cholesterol; BMI, body mass index; SBP, systolic blood pressure; DBP, diastolic blood pressure.

### Subgroup Analyses

Benefits in VO_2peak_ were directionally consistent across all prespecified strata, with no evidence of effect modification for the primary outcome (nonsignificant interaction *P* values). Significant improvements were observed in both younger and older participants, in men, in the Beijing center, and among never smokers (Figure 5). Similarly, BP effects were comparable in magnitude across strata and aligned with the overall intervention estimates at 3 months (Table S3 and S4). Office DBP showed a modest difference between high-normal and high BP strata, but other BP outcomes didn’t show the same pattern, suggesting this could be chance and should be read with caution (Table S3).

**Figure 5.**
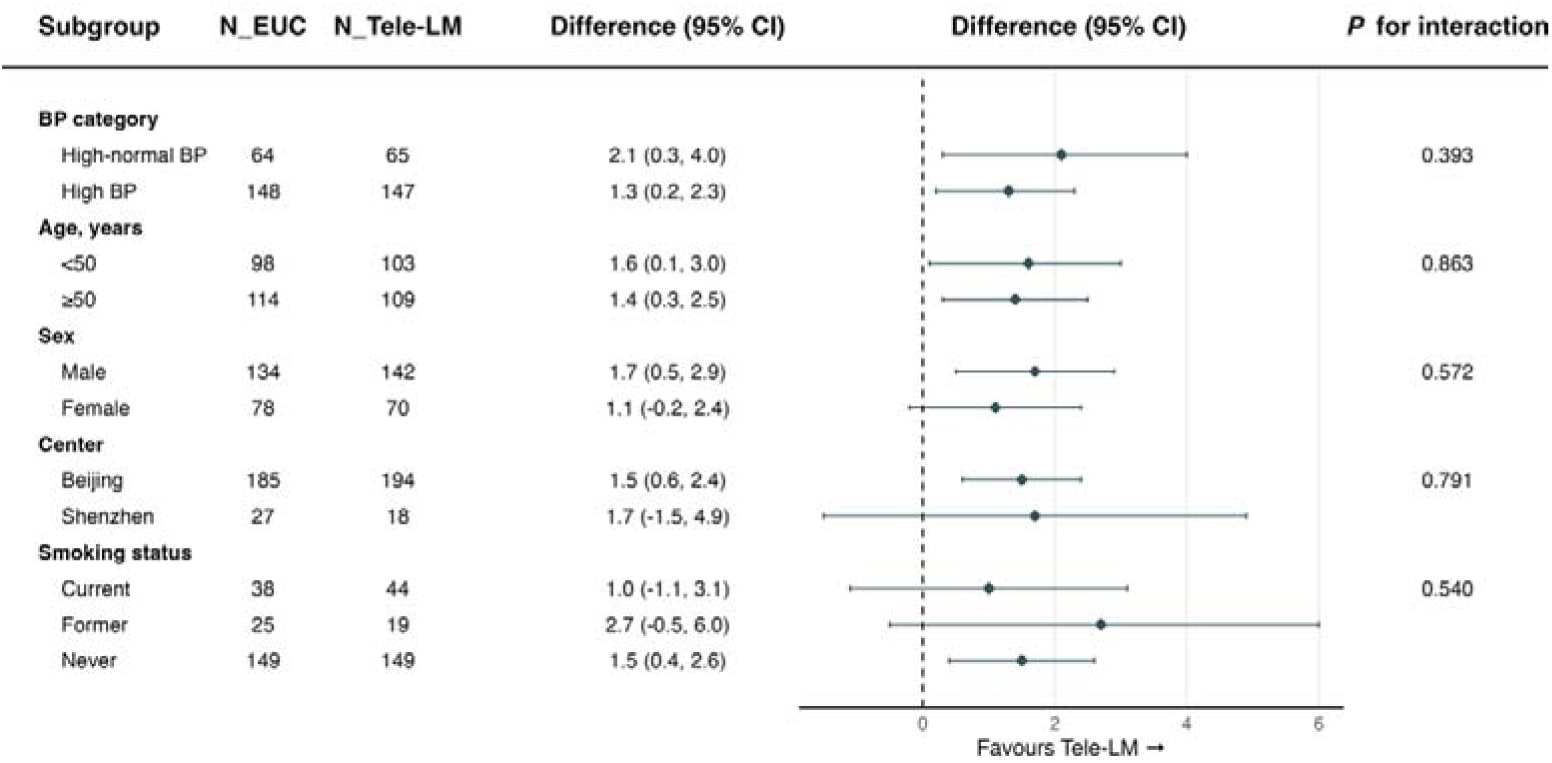
Intervention Effect on VO_2peak_ at 3 Months by Randomization Strata and Key Subgroups. EUC indicates enhanced usual care; Tele-LM, telehealth-based lifestyle modification program; VO_2peak_, peak oxygen uptake; BP, blood pressure.

### Medication Use

At study completion, a greater proportion of participants in the Tele-LM maintained or reduced their antihypertensive regimen compared with EUC (67.6% vs 53.3%). After adjustment for the baseline number of agents, the Tele-LM required fewer medications on average (adjusted mean difference –0.27 agents [95% CI, –0.44 to –0.10; *P*=0.002]). These findings indicate that the BP improvements with Tele-LM were not attributable to greater pharmacologic intensification, aligning with the study’s lifestyle-first treatment strategy.

### Safety and COVID-19 Infection

A total of 7 participants (3.3%) in the EUC and 9 (4.2%) in the Tele-LM reported AEs. The overall incidence of AEs was similar between groups, and most AEs were mild and resolved without sequelae (Table S5). None of these were considered to be related to the interventions. No serious adverse events (SAEs) were reported.

During the trial period, 328 participants (76.4% in the EUC and 78.3% in the Tele-LM) were diagnosed with COVID-19, confirmed by self-report or medical records. Most cases were mild to moderate, and no participant required hospitalization. Given the potential impact of COVID-19 on CRF and follow-up adherence, these events were monitored separately from AEs.

### Adherence

Using the predefined criterion (meeting the weekly target in ≥60% of weeks), (135/189) 71.4% of the participants in Tele-LM achieved the aerobic prescription (≥150 min/week) during months 0–3 and (81/184) 44.0% during months 4–9. When applying the full prescription (≥150 min/week of aerobic activity plus ≥2 resistance sessions/week), the corresponding proportions were (85/189) 45.0% and (43/184) 23.4%, respectively. Diet logging (uploading records ≥2 days/week in ≥60% of weeks) was (139/189) 73.5% in the first 3 months and declined to (24/184) 13.0% in months 4–9. For home BP monitoring, 78.3% of Tele-LM participants measured BP on at least 3 days per week in the first 3 months and 66.2% in the subsequent 6 months, compared with 75.6% and 66.7% in EUC, respectively.

### Sensitivity Analyses

Results were consistent across multiple imputation (MI) and CC analyses, with effect estimates similar in direction and significance. In the PP analysis, which excluded participants with poor adherence during the first 3 months, benefits of Tele-LM over EUC were more pronounced across most endpoints. For example, the adjusted mean difference in VO_2peak_ at 3 months was 1.61 mL/kg/min (95% CI, 0.71 to 2.51; *P*<0.001), exceeding the more conservative estimates from the ITT datasets. Greater improvements were also observed in CRF, home BP, office BP, and lifestyle indicators such as MVPA and DASH score. Overall, the concordance between MI and CC analyses supports the robustness of the primary findings, while PP analyses underscores the importance of adherence in maximizing intervention benefits (Figure S1–S6).

## Discussion

In this two-center randomized trial of adults with high-normal or high BP, a telehealth lifestyle program produced modest but statistically significant improvements in CRF and BP compared with EUC. Tele-LM yielded larger improvements in VO_2peak_ than EUC, surpassing it by 1.5 mL/kg/min at 3 months and 1.0 mL/kg/min at 9 months, whereas VO_2peak_ in EUC remained essentially declined slightly. Subgroup analyses revealed no significant heterogeneity for VO_2peak_ across BP category strata. Tele-LM produced greater 3-month reductions in BP by 2–4 mmHg across home and ambulatory measures, with smaller or nonsignificant differences for office SBP and nighttime DBP; effects attenuated by 9 months. Diet quality (DASH score) and MVPA increased more with Tele-LM, while weight, glycemia and lipids did not differ between groups. Sensitivity analyses including CC, MI and PP sets showed concordant effect directions, with larger estimates in the PP analysis, reinforcing robustness and highlighting the importance of adherence.

Although the absolute gain in VO_2peak_ was modest, increases of this magnitude (even 1 mL/kg/min) have been associated with meaningful reductions in cardiovascular risk,^26,27^ supporting clinical relevance. Our findings align with previous home-based or digitally guided rehabilitation trials, which have reported VO_2peak_ improvements of roughly 1–2 mL/kg/min at 6–12 months,^28^ and extend those benefits to individuals with elevated BP outside of formal cardiac rehabilitation settings. The trial coincided with the COVID-19 pandemic, during which persistent fatigue and reduced exercise tolerance have been documented even after mild infection.^29,30^ Such factors likely contributed to the slightly decline in VO_2peak_ observed in EUC. By contrast, participants in Tele-LM maintained and modestly improved CRF. This suggests that structured remote support can preserve fitness during periods of disruption, which is clinically important given the prognostic value of preventing CRF decline in at-risk populations.

Tele-LM reduced home and 24-hour SBP by about 3 mmHg more than EUC, which was smaller than that seen in some center-based, higher-intensity programs (about −7 mmHg in resistant hypertension),^8^ but is compatible with pooled effects of behavioral counseling interventions (SBP −1.8 mmHg, DBP −1.2 mmHg at 12–24 months).^31^ The comparator in this trial was active: EUC participants received home BP monitors and standardized education, measures that likely improved adherence and BP control in this arm^32^ and thereby attenuated the incremental effect of Tele-LM. Prior studies shows that self-monitoring alone can reduce BP.^33,34^ For example, Pletcher et al.^33^ found no additional SBP benefit from a connected app versus standard self-measured BP at 6 months, with both groups improving by about 11 mmHg. Taken together, these data indicate that comparators that include BP monitoring can substantially narrow between-group differences. Achieving larger incremental gains may require more intensive lifestyle counseling, clinician-supported medication titration, and stronger adherence supports. Beyond effect size, the capacity to preserve CRF and manage BP remotely at scale is clinically valuable, particularly when access to in-person care is limited, such as during the COVID-19 pandemic.^4^

Tele-LM produced early improvements in DASH score and MVPA, which diminished by 9 months as participants transitioned to self-management; walking activity and device-based step counts did not differ between groups. The absence of between-group differences in weight, glucose, and lipids likely reflects well-controlled of these parameters at baselines, a 3-month intensive phase that may be too brief for metabolic remodeling, and the pragmatic design (behavioral coaching without mandated caloric restriction or pharmacotherapy adjustment). Extending duration or adding targeted nutrition and weight-management components could plausibly yield metabolic benefit.

Effect size analyses demonstrated broadly concordant benefits of the intervention. SMD indicated small-to-moderate improvements observed across CRF, BP outcomes, and lifestyle, with negligible effects on metabolic and anthropometric measures. Most effects attenuated by 9 months. Engagement with Tele-LM was generally high during the early phase but declined over time. This trajectory was also noted in other digital lifestyle trials.^16^ In parallel, between-group differences in DASH score and MVPA diminished, and ITT estimates were smaller than those observed in PP analyses. Taken together, these patterns suggest that adherence likely contributed to the observed benefits, although formal mediation was not prespecified. From a clinical perspective, these findings highlight both the promise and the limitation of digital interventions. While they can effectively initiate behavioral change, sustaining adherence remains challenging and is essential for lasting cardiovascular benefit. The relatively low-intensity design of Tele-LM prioritized participant flexibility but limited its impact on CRF and BP. Future work should focus on remote models that combine flexibility with adherence-enhancing strategies to sustain behavioral gains and improve long-term outcomes.

The primary outcome was consistent across subgroups, with no significant heterogeneity detected. The numerically larger VO_2peak_ gains in high-normal BP may indicate greater behavioral modifiability at early disease stage or fewer comorbid constraints on training response. However, this pattern was exploratory because interaction testing was negative. Point estimates for BP reductions were somewhat larger in women, participants aged ≥50 years, those with high BP, and never smokers, but formal interaction tests were nonsignificant except for office DBP by BP category. This isolated interaction should be interpreted cautiously given the multiple comparisons and limited power. Clinically, greater BP reductions in participants with high BP likely reflect higher baseline values and greater potential for improvement,^35^ while apparent differences by age or sex may relate to behavioral, physiologic, or care-delivery factors that warrant further study. Importantly, more participants in the Tele-LM maintained or reduced antihypertensive therapy, suggesting that modest BP improvements were achieved with less pharmacologic intensification. Because medication changes were clinician-directed and not randomized, these observations remain exploratory. Future studies should prospectively assess the potential of lifestyle interventions to reduce medication requirements, ideally through standardized titration or deprescribing protocols and mediation analyses.^36^

### Strength

This trial has several notable strengths. First, it targeted individuals with high-normal or high BP, which represented a large at-risk population often overlooked in interventional research. Second, the Tele-LM integrated multiple evidence-based digital components,^37,38^ including dietary photo logging with dietitian’s feedback, automated generation of individualized aerobic and resistance prescriptions, embedded instructional videos, wearable-derived data, rule-based alerts, and motivational interviewing triggered by adherence thresholds. Together, these elements provided a coherent digital therapeutic approach grounded in established behavior changing principles. Third, the fully online and digitally automated workflow enabled uninterrupted intervention delivery during the COVID-19 pandemic, minimizing disruptions through remote coaching and Bluetooth-enabled data capture. With automating reminders, alerts, and adherence tracking, it reduced clinician workload while maintaining real-time monitoring and data-informed adjustments, thereby enhancing scalability and feasibility in real-world practice.

### Limitations

This study also has several limitations. Foremost, generalizability may be restricted, as participants were recruited from two urban centers and were comfortable using smartphones. Thus, findings may not extend to older adults, rural populations, or those with limited digital access. Second, the trial was powered primarily for VO_2peak_, so some secondary endpoints and subgroup contrasts were underpowered, and multiple comparisons raise the possibility of chance findings despite consistent effect directions. Third, MVPA was based on self-report (IPAQ) and thus prone to misclassification. Fourth, EUC included home BP monitoring and education, which may have contributed to improved outcomes and reduced between-group contrasts, although it increased pragmatic relevance. Finally, longer follow-up is needed to determine durability of effects and potential impacts on metabolic endpoints.

## Conclusion

In this randomized trial, a comprehensive telehealth-based lifestyle modification produced significant improvements in CRF and modest reductions in BP among individuals with high-normal or high BP. These benefits were achieved with minimal in-person contact and reduced reliance on pharmacologic intensification. Scalable digital approaches may complement routine hypertension care and expand the access to preventive services.

## Supporting information

protocol

## Data Availability

The data that support the findings of this study are available from the corresponding author upon reasonable request. The data are not publicly available due to ethical and privacy restrictions involving human participants.

## Acknowledgments

We thank the study participants for their invaluable contributions. We acknowledge the clinical coordinators, data managers, and research staff at both study sites for their dedicated support in recruitment, follow-up, and data collection. We particularly thank Profs. Xi Li from the National Center for Cardiovascular Diseases, Chinese Academy of Medical Sciences and Peking Union Medical College, Beijing, China, for providing expert consultation and insightful comments that substantially improved the manuscript.

## Author Contributions

The corresponding author (X.F.) had full access to all data and took responsibility for the integrity of the data and for the accuracy of the data analysis. H.D. and S.Z. formulated the study concept and design. X.S., B.W., J.D., S.Z., L.D., P.W. and M.D. carried out data acquisition. M.D., X.S., B.W., J.D., S.Z. and X.Y. carried out data analysis and interpretation. H.D. and X.S. drafted the manuscript. H.D. and M.T. reviewed and made critical revision for the manuscript. H.D. and S.Z. carried out statistical analysis. X.F. obtained funding. Y.Z., Q.X. and Z.Z. provided administrative, technical or material support. H.D. and X.Y. carried out study supervision. All authors commented on drafts of this paper.

## Nonstandard Abbreviations and Acronyms

BP: blood pressure
CRF: cardiorespiratory fitness
Tele-LM: telehealth-based lifestyle modification program
EUC: enhanced usual care
VO_2peak_: peak oxygen uptake
CPET: cardiopulmonary exercise testing
SBP: systolic blood pressure
DBP: diastolic blood pressure
FFQ: food frequency questionnaire
DASH: Dietary Approaches to Stop Hypertension
RER: respiratory exchange ratio
ABPM: ambulatory blood pressure monitoring
BMI: body mass index
LDL-C: low-density lipoprotein cholesterol
HDL-C: high-density lipoprotein cholesterol
IPAQ-SF: International Physical Activity Questionnaire–Short Form
MET: metabolic equivalent task
MVPA: moderate to vigorous physical activity
AEs: adverse events
HRmax: maximal heart rate
ITT: intention-to-treat
SD: standard deviation
CIs: confidence intervals
SMDs: standardized mean differences
CC: complete-case
PP: per-protocol
SAEs: serious adverse events
MI: multiple imputation.

---

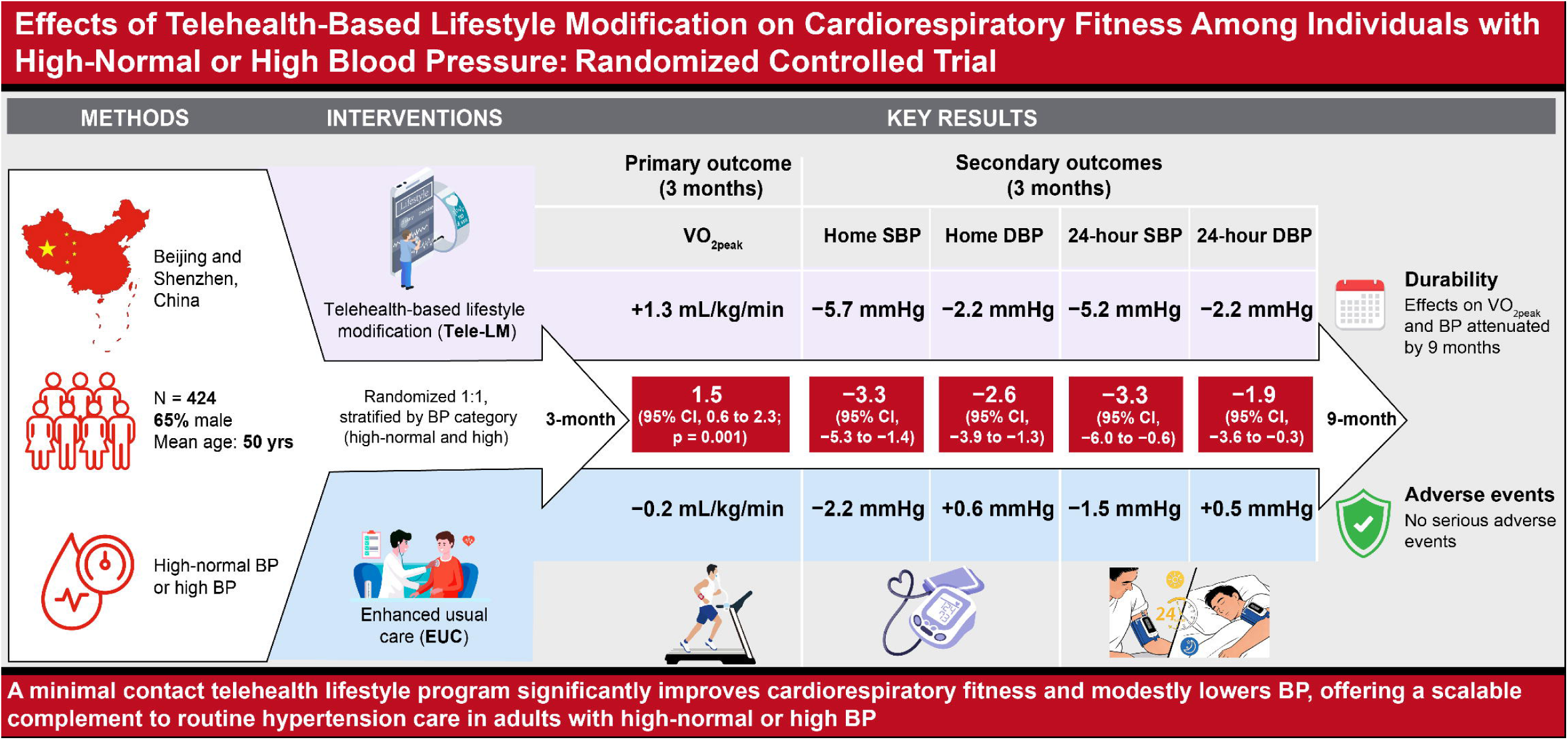

