## Supplementary material for "Effects of Telehealth-based Lifestyle Modification on Cardiorespiratory Fitness Among Individuals with High-normal or High Blood Pressure: Randomized Controlled Trial": protocol

**Supplemental Material**

**PROTOCOL**

Protocol Version: V1.5; Version Date: November 7, 2022.

Original Protocol Date: October 22, 2021.

Trial Period: July 2022–September 2024 (updated to match Methods).

Principal Investigator: Xue Feng, Center for Lifestyle Medicine, Fuwai Hospital.

Sites: Fuwai Hospital (Beijing) and Fuwai Shenzhen Hospital (Shenzhen).

Ethics: Approved by the Institutional Review Boards of both centers; written informed consent was obtained from all participants.

Reporting guideline: CONSORT.

**Background**

With socioeconomic development, lifestyle changes, population aging, and accelerated urbanization, the prevalence and mortality of cardiovascular disease (CVD) in China continue to rise. According to the Chinese Cardiovascular Health and Disease Report 2019, an estimated 245 million adults have hypertension.^1^ Serious complications of hypertension, including stroke, coronary heart disease, heart failure, and kidney disease, carry high risks of disability and death, imposing heavy burden on families and society. Evidence indicates that a reduction of 10 mmHg in systolic blood pressure (SBP) or 5 mmHg in diastolic blood pressure (DBP) is associated with an approximately 10%–15% lower risk of death, 35% for stroke, 20% for coronary heart disease, and 40% for heart failure.^2^ Therefore, effective prevention and control of hypertension is critical for curbing the CVD epidemic.

In 2003, the Seventh Report of the Joint National Committee (JNC 7) introduced the concept “high-normal blood pressure (BP),” describing the condition when someone who is not on antihypertensive medication having seated SBP between 120–139 mmHg and/or DBP between 80–89 mmHg on at least two occasions.^3^ High-normal BP is a transitional stage from normal BP to hypertensive status. This condition is a prodrome that significantly elevates the risk for clinical hypertension and subsequent CVDs. Elevated BP results from interactions between genetic and environmental factors, including heredity, age, overweight/obesity, high sodium intake, smoking, excess alcohol, insufficient physical activity, and chronic stress.

The greater the number and severity of individual risk factors, the higher the BP and the greater the risk of developing hypertension.^4,5^ To delay progression to hypertension and mitigate associated cardiovascular and cerebrovascular risks, lifestyle modification is strongly recommended.^3^ Key detrimental factors include excessive intakes of sodium, fat and alcohol, with meta-analyses confirming that reducing salt intake effectively lowers BP.^6,7^ Similarly, smoking elevates BP and heart rate, with smokers demonstrating higher SBP and DBP than in non-smokers; cessation is critical for reducing CVD progression in hypertensive patients and lowering long-term mortality among patients with coronary heart disease.^8^ Moreover, obesity triples the risk of hypertension compared to a normal body mass index (BMI).^9^ Conversely, regular physical activity significantly reduces hypertension risk while improving overall fitness and health. Hypertension is also strongly associated with chronic psychological stress, anxiety, and high pressure. Prior studies indicate substantial prevalence of anxiety and depression among patients with hypertension,^10^ and these symptoms can impede treatment response. Therefore, comprehensive lifestyle management encompassing dietary change, smoking and alcohol cessation, adequate exercise, and psychological balance is fundamental to improve clinical outcomes. The integrated approach aids in controlling BP, enhancing treatment efficacy, and preventing cardiovascular events in patients with hypertension or high-normal BP.

Cardiorespiratory fitness (CRF) is closely associated with progression of hypertension. Lower CRF is related to faster progression and higher risk of adverse cardiovascular events.^11,12^ Individuals with poor CRF have higher BP than fit individuals^13^. As an objective physiologic indicator of habitual physical activity, CRF is strongly associated with CVD and all-cause mortality across healthy individuals and those with hypertension or other CVDs.^14,15^ Cross-sectional data further show relationships between CRF and unhealthy lifestyle (physical inactivity, smoking) and with chronic diseases (obesity, dyslipidemia, hypertension, diabetes).^16^ Thus, improving CRF is an important component in BP management. Prior randomized controlled trials (RCTs) suggest that lifestyle management can improve CRF among patients with hypertension,^17^ supporting the potential benefits of comprehensive lifestyle intervention.

Cardiopulmonary exercise testing (CPET) provides an objective and quantitative assessment of CRF and functional status in hypertensive and high-normal BP individuals. It is a noninvasive test that evaluates cardiopulmonary and musculoskeletal responses during exercise, hemodynamic and oxygen kinetics metrics, cardiopulmonary function, and exercise tolerance.^18^

Long-term BP assessment requires extended monitoring, which is often challenging to obtain in community clinics or hospitals. Ambulatory and wearable monitoring can facilitate data collection and mitigate white-coat effects. Meanwhile, lifestyle habits take longer time to change. Poor adherence and lack of tailored guidance can limit outcomes. Accordingly, individualized, specific, and stepwise comprehensive interventions are needed. With advances in artificial intelligence and internet technology, digital platforms have been used to manage health, such as wearable devices for physical activity and weight control,^19^ remote smartphone monitoring of heart failure for self-care and clinical management,^20^ and telemedicine-supported systems for early type 2 diabetes management.^21^ Several trials have shown that home‑based or tele‑monitored exercise programs can modestly enhance CRF and reduce BP.^22,23^ A recent small study in the US found that mobile device–based home BP monitoring and management among hypertensive patients was feasible and effective to reduce BP.^24^ However, hypertension apps co-developed with healthcare professionals remain uncommon. Few products have undergone rigorous clinical evaluation, and findings for blood pressure control are inconsistent. Notably, no studies have focused specifically on adults with high‑normal or well‑controlled hypertension to determine whether telehealth‑delivered lifestyle modification further enhances CRF and yield additional cardiovascular benefits.

To address this gap, we design a two‑center, stratified, randomized controlled trial to evaluate the effect of a comprehensive lifestyle modification program delivered through a mobile application for individuals with high‑normal or high BP. The intervention combines personalized dietary coaching, supervised aerobic and resistance exercise prescriptions, continuous BP self‑monitoring, and motivational interviewing, all supported by wearable devices and a smartphone platform. We hypothesize that this telehealth‑based approach will elicit clinically meaningful improvements in CRF, augment BP control, and promote healthier lifestyle behaviors.

**Objectives**

This study will utilize smart wearable devices to dynamically monitor multiple physiologic parameters in adults with high BP or high-normal BP. Based on continuous collected data, personalized lifestyle guidance will be provided. We aim to evaluate the effectiveness of a telehealth‑based lifestyle motification system in improving cardiorespiratory fitness and other physiological indicators. The ultimate goal is to explore the feasibility of applying a telehealth-based system for chronic-disease management, and to provide an evidence-based foundation for effective and early intervention for hypertension.

**Methods**

**Trial Design**

This is a prospective, two-center, stratified, randomized, controlled, single-blind trial conducted in urban clinics in China between July 2022 and September 2024 (updated to match Methods). Randomization is stratified by BP category (untreated high-normal BP and high BP) and assigned 1:1 to Telehealth-based Lifestyle Modification (Tele-LM) or Enhanced Usual Care (EUC) using a computer-generated randomization sequence. Allocation concealment will use sealed, opaque envelopes. Outcome assessors are blinded while participants and staff who perform interventions are not.

**Participants**

Setting and recruitment: Consecutive sampling will take place at Fuwai Hospital (Beijing) and Fuwai Shenzhen Hospital from 2022 to 2023, recruiting adults diagnosed with high BP (SBP ≥140 mmHg and/or DBP ≥90 mmHg, or on antihypertensive therapy) or high-normal BP (SBP 120–139 mmHg and/or DBP 80–89 mmHg) who volunteer to participate.

Inclusion criteria (all required): (1) age ≥18 and <70 years; (2) having high BP or high-normal BP as defined above; (3) ownership and competence in using an internet-enabled smartphone (Android preferred); (4) signed informed consent.

Exclusion criteria (any): (1) acute myocardial infarction, tachyarrhythmia, pulmonary edema, severe aortic stenosis, or other serious cardiorespiratory disorders; (2) acute phase of cardiovascular or cerebrovascular disease; (3) poorly controlled hypertension (>160/100 mmHg despite treatment); (4) conditions preventing CPET such as fracture or unstable joints; (5) skin disease or injury at device application sites; (6) active psychiatric illness, epilepsy, or other conditions causing involuntary movement; (7) pacemaker implantation, pregnancy or intention to become pregnant, or severe allergy/atopy.

**Trial Arms and Procedures**

Participants currently on antihypertensive medications are encouraged to continue under the guidance of their physicians.

***Control: Enhanced Usual Care (EUC)***

Participants randomized into EUC group will receive the following intervention components: usual clinical care plus onsite lifestyle education; monitoring with dynamic electrocardiogram (ECG), home BP, ambulatory BP monitoring (ABPM), and use of smartwatch for activity and sleep tracking. Participants will have access to educational materials via the Healthy Lifestyle app for a total of nine months to support self-management and data uploads. Automated alerts are triggered for excessively high (>180/110 mmHg) or low (SBP <100 mmHg) home BP values, prompting timely medical consultation.

***Intervention: Telehealth-based Lifestyle Modification (Tele-LM)***

Participants randomized into Tele-LM group will receive the following intervention components: a comprehensive multi-component telehealth program delivered through the Healthy Lifestyle App (OPPO Mobile Telecommunications Co., Guangdong, China). Participants will have access to personalized lifestyle prescriptions, tracking reminders, educational materials, monitoring, feedback, self-management tools, and communication through this platform. Recommendations are tailored using inputs on personality traits, behavioral characteristics, and hypertension determinants.

Dietary management

At enrollment, dietitians will provide healthy eating education and personalized diet prescriptions through the app based on semi-quantitative food frequency questionnaire (FFQ) inputs and Diet Approaches to Stop Hypertension (DASH) dietary principles. Diet prescriptions include calorie goal, dietary structure, preferred foods, and recommendations to reduce salt and alcohol. Participants will be instructed to upload meal photos. Dietitians will provide weekly feedback regarding intakes of fruits, vegetables, whole grains, sodium, saturated fat, and overall DASH adherence.

Exercise management

Individualized aerobic and resistance exercise prescriptions will be generated automatically based on baseline CPET results and participant preferences, targeting at least 150 minutes per week of moderate-intensity aerobic exercise plus two resistance sessions per week. Resistance training instructional videos will be provided in-app. Smartwatch (OWW211 OPPO Watch 3 Pro) is used to record heart rate and steps. The app logged adherence and prompted sessions. Automated alerts will be generated if heart rate exceeds predefined safety threshold. Physical therapists will provide weekly feedback, with adjustments as needed.

BP monitoring and feedback

Home BP monitoring: Participants will receive validated BP monitor (U36T; Omron Healthcare Co., China) to measure BP in the mornings and evenings of at least three days per week. Readings are synced via Bluetooth to the app for remote review. Alerts will be generated with markedly elevated (>180/110 mmHg) or abnormally low (SBP <100 mmHg), prompting participants to seek care within three days.

Motivational interviewing

If adherence to dietary or exercise prescriptions falls below 50%, a motivational interviewing by clinical psychologists will be conducted via phone or video to strengthen behavior change and problem solving.

Tele-LM participants will need actively engage in for three months, followed by six months of self-management (month 4–9) with ongoing self-monitoring via the app.

**Baseline Assessments**

Demographics (sex, age, occupation, education, household income per-capita), smoking and alcohol history, allergy history, family history of chronic disease, lifestyle (diet, physical activity, sleep), and medication history will be collected via on-site guidance. Data will be entered through the Research app and imported into the case report form (CRF).

***Physiologic measurements:***

Anthropometrics: Height, weight, circumferences measured by calibrated instruments will be recorded in the CRF.

Office BP will be measured at each visit. Two readings will be taken per visit, and the average will be recorded if the difference is <10 mmHg. If the difference exceeds 10 mmHg, additional readings need to be obtained until two stable values are achieved.

***Cardiopulmonary exercise testing (CPET):***

Cycle ergometer with a ramp protocol; oxygen consumption (VO₂), carbon dioxide production (VCO₂), peak oxygen uptake (VO_2peak_), respiratory rate, tidal volume; concurrent ECG, oxygen saturation in blood (SpO₂), and BP monitoring (MasterScreen® cycle ergometer, Vyaire Medical, Hoechberg, Germany) will be assessed and reported.

Physical fitness testing: Upper- and lower-limb muscle strength (arm-curl test; chair-stand test), balance, flexibility (sit-and-reach; back scratch), will be measured by trained staff.

***Laboratory tests:***

Fasting blood analyses including glucose, lipids, free fatty acids, and selected nutritional markers (e.g., 25(OH)D, magnesium) will be measured. Participants will be asked to fast for at least eight hours prior to blood draw; About 15 mL venous blood will be drawn and analyzed on automated platforms.

***Sleep quality:***

Pittsburgh Sleep Quality Index (PSQI) will be used to reflect sleep quality.

***Quality of life (QoL) and psychosocial health:***

Short Form-36 (SF-36), Patient Health Questionnaire-9 (PHQ-9), and the 7-item Generalized Anxiety Disorder 7 (GAD-7) will be used to measure QoL, depression, and anxiety, respectively.

Nutrition assessment: A semi-quantitative FFQ (SQFFQ) will be conducted by the dietitians to evaluate dietary patterns over a period of three months.

**Intervention Period (Months 0–3)**

Tele-LM participants will receive a three-month lifestyle intervention and dynamic monitoring. EUC participants will receive dynamic monitoring only.

***Lifestyle intervention:***

Lifestyle intervention components and frequencies are summarized in Table 1.

**Table 1. Intervention components and frequencies of Tele-LM group**

| **Module** | **Intervention Content** | **Frequency** |
| --- | --- | --- |
| **Dietary management** | Healthy eating counseling;  Individualized diet prescription entered in the Research app;  Meal photos upload | Counseling at enrollment;  Feedback upon photo uploads;  Biweekly phone follow-ups. |
| **Exercise management** | Aerobic and resistance training plan with individualized targets based on status and CPET to define heart rate zones, repetitions, and duration;  App push notifications of exercise tasks | Plan adjusted every 2 weeks according to participant’s execution;  Exercise supervision and feedback. |
| **Psychological balance** | Motivational interviewing;  App push notifications of mindfulness/meditation videos;  Periodic QoL and psychosocial questionnaires. | One motivational interviewing session at enrollment;  Additional motivational interviewing sessions every 2 weeks based on diet/exercise adherence. |
| **Smoking/alcohol education** | Health-education articles and reminders for risk-factor control. | Irregular push notifications. |

***Dynamic monitoring devices and schedules:***

Dynamic ECG: Single-lead long-term recorder (24 h weekly) ECG device will be used to screen arrhythmia with day/night stratification. Data is limited to research analysis only. It will not be utilized to adjust exercise prescriptions.

Ambulatory BP monitoring (ABPM): An ABPM device (M2; Raycome Health Technology, Shenzhen, China) scheduled to collect data hourly while awake (06:00–22:00) and every 2 h during sleep (22:00–06:00) will be provided to all participants. A session is considered valid if there is ≥70% of scheduled readings. Otherwise, participants will need to repeat the next day. Derived metrics include 24-h/daytime/nighttime means, BP load, and dipping ratio ((day − night)/day ×100%).

Home BP monitoring: An upper-arm electronic device (U36T; Omron) is recommended to use for BP monitoring twice daily (morning, evening), 3–5 days per week except on ABPM days; Measurements will be done when participants are seated and rest for at least 5 minutes. These readings supplement the BP tracking during non-ABPM periods.

Smartwatch monitoring: OPPO Watch (OWW211 OPPO Watch 3 Pro) capturing steps, activity energy, exercise duration and frequency will be provided. The PPG/ECG sensors provide data for heart rate (HR), SpO₂, arrhythmia screening, sleep tracking and Heart rate variability (HRV)-based stress.

**Follow-up (Months 4–9)**

Both groups will be followed up to Month 9. Dynamic ECG, home BP, and smartwatch monitoring are still required for all participants during follow-up periods. Lifestyle coaching is no longer proactively provided. Participants can choose to maintain lifestyle behaviors developed during the intervention.

Clinic assessments will occur at months 3, 6, and 9, including assessments of CPET (Months 3 and 9), ABPM, anthropometrics, clinical and laboratory tests, physical fitness, and psychosocial questionnaires, which are consistent with baseline procedures. Table 2 demonstrates all the clinic assessment items and their corresponding frequencies.

**Table 2. Schedule of Assessments**

| **Assessment** | **Baseline (Day 0)** | **Month 3** | **Month 6** | **Month 9** |
| --- | --- | --- | --- | --- |
| Eligibility & Consent | X |  |  |  |
| Randomization & Device Training | X |  |  |  |
| Demographics/Lifestyle/History | X |  |  |  |
| Medication Use | X | X | X | X |
| Anthropometrics | X | X | X | X |
| Laboratory tests | X | X |  | X |
| CPET | X | X |  | X |
| Physical fitness tests | X | X | X | X |
| Nutrition assessment (FFQ/DASH score) | X | X | X | X |
| Psychosocial (SF-36/PHQ-9/GAD-7/PSQI) | X | X | X | X |
| Dynamic ECG |  | X | X | X |
| ABPM |  | X | X | X |
| Home BP |  | X | X | X |
| Smartwatch metrics |  | X | X | X |
| Adverse events |  | X | X | X |

**Impact of COVID-19 on Study Procedures (Protocol Adaptation)**

In December 2022, new enrollments and in-person data collection were halted due to the COVID-19 pandemic. This led to subsequent adjustments to the study protocol to maximize data collection from enrolled participants. Specifically, the follow-up window for outcome assessments was extended, and individualized modifications were made to the exercise prescriptions to accommodate participants recovering from COVID-19 symptoms. Participants were permitted to resume the Tele-LM program upon receiving medical clearance.

Participants who missed the 3-month clinical assessments due to COVID-related illness or restrictions were permitted to complete these assessments during the 6-month visit. These changes were implemented consistently across study centers and documented to ensure accurate interpretation of results. Nonetheless, the pandemic increased the risk of missing data during follow-up.

**Outcomes**

***Primary outcome:***

The primary outcome is the change of VO₂peak (mL/kg/min) by CPET from baseline to month 3.

***Secondary outcomes:***

The secondary outcomes include the following parameters:

- Other CPET parameters and changes: anaerobic threshold, VO₂, VCO₂, respiratory rate, tidal volume, SpO₂; HRmax, peak workload, and RER at 3 and 9 months;
- Body weight and BMI;
- ECG and BP (home, ABPM, and office), and their changes;
- Laboratory indices and changes: fasting glucose and lipids, etc;
- Physical fitness: muscle strength, balance, flexibility and their changes;
- Wearable-derived key indices (e.g., SpO₂, arrhythmia flags), daily activity averages, and improvements across time segments;
- Sleep quality (PSQI), quality of life (SF-36), and mental health scores (PHQ-9, GAD-7);
- Adverse events: start/end time, frequency, symptoms, severity, actions taken, and outcomes.
- Adherence measures: include adherence to lifestyle management delivered via the Research app, completion and target attainment for online lifestyle prescriptions, changes in smoking/alcohol use, device-use adherence and user satisfaction.

**Sample Size**

Sample size estimation was based on previous trials.^25,26^ Stratifying by BP category, we assumed between-group differences of VO_2_peak after 3 months were 2.0±4.5 for people with high BP, and 3.0±4.5 for people with high-normal BP. With Bonferroni-adjusted two-sided α=0.025 and 90% power, the required sample sizes were 124 for hypertension and 52 for high-normal BP, which meant a total of 176 participants per arm. Allowing for a 20% attrition, the target sample size was 212 per arm. A total of 424 participants is required. Recruitment within a stratum will be terminated once its target is reached.

**Statistical Analysis Plan**

Analyses follow the intention-to-treat (ITT) principle. If the 3-month primary outcome is missing but the 6-month value is available, the 6-month value is substituted. Remaining missing data are handled by multiple imputation with chained equations (100 imputations) under MAR. Continuous variables are summarized as mean±SD. Categorical variables are summarized as counts and percentages.

General linear models will used to analyze between-group changes of VO₂peak from baseline to month 3, adjusted for baseline VO₂peak, BP category, center, age, sex, education level, income, and smoking. Secondary outcomes will be analyzed similarly with baseline adjustment. Standardized mean differences are reported for all continuous outcomes. Prespecified subgroup analyses compare intervention effects across strata defined by BP category, age, sex, center, and smoking. Sensitivity analyses will be used to assess the robustness of findings by using complete-case and per-protocol analyses (excluding Tele-LM participants who achieved planned weekly exercise in <50% of weeks during months 0–3). Two-tailed P<0.05 is considered significant. Analyses will be done using R version 4.4.2 (R Foundation for Statistical Computing, Vienna, Austria). Figures will be generated using ggplot2 package.

**Data Management**

To ensure the accuracy and authenticity of data collection, the research team will conduct data quality-control training. The data in this study will be sourced from clinical examinations, wearable devices, and follow-up records collected by researchers. Clinical and wearable device data will be extracted through automated systems. A confidentiality protocol will be strictly followed in terms of data checking and export. The exported data will be stored exclusively on a computer without internet access by the researchers involved in this study. Designated research physicians will be responsible for data collection and entry. All data will be entered for twice by two researchers separately. Two sets will be compared using data comparison function of software to prevent any data errors like missing values or outliers.

**Confidentiality**

The study will strictly adhere to the confidentiality principles in accordance with regulations. The research platform employs information encryption technology and identity-based authorization controls, assigning differentiated access permissions to research team personnel. Data desensitization techniques are applied to de-identify personal information, thereby protecting private data and ensuring information security during backend collection, extraction, and analysis.

All materials will be properly preserved as research evidence and will not be used for any other purposes. Only the research team, relevant research administration department, and the ethics committee are permitted to access study-related materials. No individual identities or personal data will be disclosed in any research reports. Should any third parties require data access for analysis purpose, explicit authorization from the principal investigators must be obtained prior to any usage.

The data and results of this study may be published in print of electronically in domestic or foreign medical journals or other media. The researchers will keep the information confidential to ensure that the personal information of the research participants will not be disclosed.

**Quality Control**

***Device operation training:***

Standard Operating Procedure (SOP) will be established before the study begins. All research team members are required to get trained on the study protocol and device use/maintenance to ensure consistency.

Site staff is responsible for all onsite clinical evaluations. Site staff who perform clinical assessments should completely understand the study protocol. Site staff should be familiar with how to use all devices, maintain and troubleshooting.

The lead coordinating center (Fuwai Hospital (Beijing)) is responsible for all participants’ diet and exercise prescriptions, motivational interviewing, and adherence management.

***Process control and data recording:***

The entire process must be conducted under stringent operating procedures. The study personnel must complete all sections of the CRF accurately and thoroughly in accordance with the guidelines to ensure the integrity and authenticity of the data. All observations obtained during the study must be verified to ensure data reliability, thereby ensuring all conclusions are derived from the source data.

***Safety monitoring:***

Participants must strictly follow their exercise prescriptions formulated by the research doctor. They are required to measure BP and heart rate before, during, and after each exercise session. They are also required to carry nitroglycerin, and wear a smartwatch and a dynamic ECG monitor to record their heart rate and ECG. The most appropriate hear rate range will be reminded to ensure the effectiveness and safety of home exercise. Data will be uploaded to the platform in real time. Adverse events are defined as any adverse and unexpected clinical signs, symptoms or diseases related to the lifestyle intervention plan. The research team will evaluate the severity and its correlation with the intervention plan. Severe adverse events include falls and malignant arrhythmias during exercise.

**Ethics**

***Ethics approval:***

Protocol and informed consent will need to be approved by the ethics committee of Fuwai Hospital prior to initiation. During the implementation of the study, it is required to strictly follow all the regulations stipulated in the plan. The researchers will not modify or change the operational procedures described in the research plan. If modifications to the research plan and informed consent form are needed during the research process, they must be re-submitted to the ethics committee for approval.

***Informed consent process:***

Before each participant is included in this study, the researchers will provide a complete, comprehensive, easy-to-understand, and ethically approved informed consent document and its explanations in written form to the participants or their legal representative, and give them or their legal representative sufficient time to consider whether to participate in this study. The research can only proceed after the participant or their legal representative sign the informed consent document.

During any stage of the study, the participants have the right to withdraw from the study at any time without facing discrimination or retaliation. Their medical treatment and rights will not be affected, and they can still receive other treatment methods. During the study period, if new safety information leads to a significant change in the risk/benefit assessment, all updated new information should be provided to the subject or their legal representative, and a new informed consent document should be re-signed.

***Risk minimization:***

This study will fully consider the informed consent and interests of the participants. The monitoring by the wearable devices is a non-invasive examination. Currently, there have been no reports of any adverse events. The study requires continuous wearing of these devices, which may cause pressure on the tested limbs and discomfort for the participants. This can be avoided by adjusting the appropriate tightness and wearing position. The home exercise intervention in the intervention group will be based on the appropriate exercise prescription and target heart rate assessed by clinical evaluation at baseline. The smartwatch will provide heart rate monitoring prompts to ensure the effectiveness and safety of home exercise.

All the operations in this study are routine clinical operations, and will not cause any additional risks or harm to the participants. The researchers will provide equipment operation training to the participants to ensure that no errors occur during use.

The intervention group may have better benefits than the control group, or it may be ineffective. Completing this study will be beneficial to promoting the development process of smart Internet of Things and smart wearable devices, and improving the convenience of lifestyle intervention for hypertensive patients.

**Study Timeline**

The total duration of this study is estimated to be approximately 24 months based on the number of cases, informed consent, and the screening and enrollment process. If there is difficulty in actual operations, the duration may be extended to meet the sample size and clinical research content requirements.

**Organization and Management**

This work is supported by OPPO Mobile Telecommunications Co. (Guangdong, China). OPPO Co. provides financial support, study equipment, and technical resources but has no role in the design of the analysis plan, data interpretation, or the decision to publish the results. Study operations team is Center for Lifestyle Medicine, Fuwai Hospital (PI: Xue Feng). Team responsibilities include recruitment, randomization, data handling, intervention delivery, follow-up, quality control, statistical analysis, and reporting.
