## Supplementary material for "Effects of Telehealth-based Lifestyle Modification on Cardiorespiratory Fitness Among Individuals with High-normal or High Blood Pressure: Randomized Controlled Trial": table S1-S5; figure S1-S6

**Supplemental Material**

### **Table S1. Scoring Criteria for DASH Diet Adherence**

| **DASH Diet Index Item** | **Score** |
| --- | --- |
| **Total Grain** |  |
| Whole grains and tubers ≥50% of all grain intake | 1 |
| Whole grains and tubers <50% of all grain intake | 0.5 |
| Rarely consume whole grains or tubers | 0 |
| **Vegetables** |  |
| ≥300 g/day | 1 |
| 200–300 g/day | 0.5 |
| <200 g/day | 0 |
| **Fruits** |  |
| ≥200 g/day | 1 |
| <200 g/day | 0.5 |
| Almost none | 0 |
| **Dairy** |  |
| ≥200 g/day | 1 |
| <200 g/day | 0.5 |
| Almost none | 0 |
| **Meat, poultry, and eggs** |  |
| ≤180 g/day | 1 |
| 180–360 g/day | 0.5 |
| >360 g/day | 0 |
| **Fish** |  |
| ≥150 g/week | 1 |
| <150 g/week | 0.5 |
| Almost none | 0 |
| **Nuts, seeds, and legumes** |  |
| ≥70 g/week | 1 |
| 35–70 g/week | 0.5 |
| <35 g/week | 0 |
| **Oil** |  |
| <25 g/day | 1 |
| 25–35 g/day | 0.5 |
| >35 g/day | 0 |
| **Sweets** |  |
| No frequent consumption of sugar-rich foods | 1 |
| Frequent consumption of one type of sugar-rich food | 0.5 |
| Frequent consumption of ≥2 types of sugar-rich foods | 0 |
| **Sodium** |  |
| Low salt use and rarely consume pickled/salted foods | 1 |
| Moderate salt use or occasional intake of pickled/salted foods | 0.5 |
| High salt use or frequent intake of pickled/salted foods | 0 |

The assessments of total grain, sweets, and sodium were based on qualitative survey responses. Sugar-rich foods included in the questionnaire were: cookies or pastries, sugar-sweetened beverages, candied or preserved fruits. Sodium intake assessment was derived from reported salt use and consumption frequency of pickled/salted foods.

### **Table S2. Mean Changes from Baseline to 3 Months and 9 Months for Primary and Secondary Outcomes in EUC and Tele-LM**

|  | **EUC (N=212)** | | **Tele-LM (N=212)** | | **Unadjusted mean difference (95%CI)** | **Adjusted mean difference (95%CI)^*^** | **Adjusted *P* for difference** |
| --- | --- | --- | --- | --- | --- | --- | --- |
|  | **Mean (SD)** | **Mean change from baseline (95%CI)** | **Mean (SD)** | **Mean change from baseline (95%CI)** |  |  |  |
| **Cardiorespiratory Fitness** | | | | | | | |
| VO_2peak_, mL/kg/min | | | | | | | |
| Baseline | 22.9 (5.7) | – | 23.0 (5.9) | – | – | – | – |
| 3 months | 22.7 (5.9) | -0.2 (-0.8, 0.4) | 24.3 (6.1) | 1.3 (0.6, 1.9) | 1.5 (0.5, 2.4) | 1.5 (0.6, 2.3) | 0.001 |
| 9 months | 22.4 (5.6) | -0.5 (-1.1, 0.1) | 23.6 (6.0) | 0.6 (-0.0, 1.3) | 1.1 (0.1, 2.1) | 1.0 (0.1, 1.9) | 0.027 |
| HRmax, bpm | | | | | | | |
| Baseline | 139.9 (22.2) | – | 143.6 (21.0) | – | – | – | – |
| 3 months | 139.5 (22.9) | -0.4 (-2.7, 1.8) | 148.7 (23.3) | 5.1 (2.6, 7.6) | 5.5 (1.8, 9.2) | 6.7 (3.1, 10.3) | <0.001 |
| 9 months | 137.7 (21.5) | -2.2 (-4.4, 0.1) | 147.2 (21.4) | 3.6 (1.3, 5.8) | 5.8 (2.3, 9.2) | 7.0 (3.7, 10.2) | <0.001 |
| Peak workload, Watt | | | | | | | |
| Baseline | 136.7 (57.9) | – | 140.8 (47.8) | – | – | – | – |
| 3 months | 135.5 (45.8) | -1.1 (-8.1, 5.8) | 151.2 (49.4) | 10.4 (6.8, 14.1) | 11.6 (3.1, 20.1) | 12.6 (6.0, 19.2) | <0.001 |
| 9 months | 134.4 (46.0) | -2.3 (-9.3, 4.7) | 149.9 (49.2) | 9.1 (5.1, 13.2) | 11.4 (2.7, 20.2) | 12.0 (5.3, 18.7) | <0.001 |
| RER | | | | | | | |
| Baseline | 1.2 (0.1) | – | 1.2 (0.1) | – | – | – | – |
| 3 months | 1.2 (0.1) | -0.0 (-0.0, 0.0) | 1.2 (0.2) | 0.0 (-0.0, 0.0) | 0.0 (-0.0, 0.1) | 0.0 (-0.0, 0.1) | 0.073 |
| 9 months | 1.2 (0.1) | -0.0 (-0.0, 0.0) | 1.2 (0.1) | -0.0 (-0.0, 0.0) | 0.0 (-0.0, 0.0) | 0.0 (-0.0, 0.0) | 0.602 |
| **Blood Pressure, mmHg** | | | | | | | |
| Home SBP |  |  |  |  |  |  |  |
| Baseline | 128.4 (12.4) | – | 128.9 (10.5) | – | – | – | – |
| 3 months | 126.1 (10.4) | -2.2 (-3.7, -0.7) | 123.2 (9.1) | -5.7 (-7.0, -4.3) | -3.4 (-5.8, -1.1) | -3.3 (-5.3, -1.4) | <0.001 |
| 9 months | 125.7 (10.8) | -2.7 (-4.4, -1.1) | 123.7 (8.9) | -5.2 (-6.8, -3.7) | -2.5 (-5.1, 0.0) | -2.2 (-4.3, -0.2) | 0.032 |
| Home DBP |  |  |  |  |  |  |  |
| Baseline | 81.7 (7.2) | – | 82.4 (6.8) | – | – | – | – |
| 3 months | 82.3 (7.1) | 0.6 (-0.4, 1.5) | 80.2 (6.8) | -2.2 (-3.1, -1.4) | -2.8 (-4.2, -1.3) | -2.6 (-3.9, -1.3) | <0.001 |
| 9 months | 81.8 (6.9) | 0.1 (-0.8, 1.1) | 79.8 (6.8) | -2.5 (-3.5, -1.5) | -2.7 (-4.3, -1.0) | -2.4 (-3.8, -1.0) | <0.001 |
| Morning home SBP | | | | | | | |
| Baseline | 127.9 (12.3) | – | 128.5 (11.1) | – | – | – | – |
| 3 months | 127.0 (11.0) | -0.9 (-2.5, 0.6) | 123.9 (9.6) | -4.7 (-6.1, -3.2) | -3.7 (-6.3, -1.1) | -3.5 (-5.7, -1.4) | 0.001 |
| 9 months | 126.7 (11.7) | -1.2 (-2.9, 0.6) | 124.0 (9.2) | -4.6 (-6.2, -2.9) | -3.4 (-6.3, -0.5) | -3.0 (-5.3, -0.7) | 0.01 |
| Morning home DBP | | | | | | | |
| Baseline | 82.5 (7.4) | – | 83.4 (7.4) | – | – | – | – |
| 3 months | 83.3 (7.4) | 0.8 (-0.2, 1.8) | 81.0 (7.1) | -2.4 (-3.3, -1.5) | -3.2 (-4.7, -1.7) | -2.9 (-4.2, -1.6) | <0.001 |
| 9 months | 82.9 (7.4) | 0.4 (-0.7, 1.5) | 80.7 (7.0) | -2.7 (-3.8, -1.7) | -3.1 (-4.9, -1.3) | -2.6 (-4.2, -1.1) | <0.001 |
| Evening home SBP | | | | | | | |
| Baseline | 129.2 (12.7) | – | 128.6 (11.3) | – | – | – | – |
| 3 months | 126.5 (11.2) | -2.7 (-4.3, -1.0) | 122.9 (9.5) | -5.7 (-7.2, -4.2) | -3.0 (-5.7, -0.4) | -3.7 (-5.7, -1.6) | <0.001 |
| 9 months | 125.5 (11.6) | -3.7 (-5.6, -1.8) | 122.4 (9.1) | -6.2 (-7.8, -4.6) | -2.5 (-5.3, 0.4) | -3.1 (-5.3, -0.8) | 0.007 |
| Evening home DBP | | | | | | | |
| Baseline | 81.8 (8.2) | – | 82.3 (7.3) | – | – | – | – |
| 3 months | 81.8 (7.4) | 0.0 (-1.1, 1.1) | 79.8 (7.0) | -2.6 (-3.5, -1.6) | -2.6 (-4.2, -0.9) | -2.5 (-3.9, -1.1) | <0.001 |
| 9 months | 81.1 (7.3) | -0.7 (-1.9, 0.5) | 78.9 (7.2) | -3.4 (-4.5, -2.4) | -2.7 (-4.6, -0.8) | -2.6 (-4.2, -1.0) | 0.001 |
| 24-hour ambulatory SBP | | | | | | | |
| Baseline | 128.8 (12.3) | – | 129.6 (10.6) | – | – | – | – |
| 3 months | 127.3 (11.6) | -1.5 (-3.3, 0.4) | 124.4 (11.0) | -5.2 (-6.9, -3.5) | -3.7 (-6.8, -0.7) | -3.3 (-6.0, -0.6) | 0.019 |
| 9 months | 128.0 (11.9) | -0.8 (-2.6, 1.0) | 126.5 (10.3) | -3.1 (-4.7, -1.4) | -2.3 (-5.0, 0.4) | -1.8 (-4.0, 0.5) | 0.122 |
| 24-hour ambulatory DBP | | | | | | | |
| Baseline | 81.2 (7.1) | – | 82.5 (6.8) | – | – | – | – |
| 3 months | 81.7 (7.5) | 0.5 (-0.6, 1.5) | 80.3 (7.2) | -2.2 (-3.3, -1.1) | -2.7 (-4.5, -0.9) | -1.9 (-3.6, -0.3) | 0.020 |
| 9 months | 81.3 (7.0) | 0.1 (-0.9, 1.1) | 80.9 (7.0) | -1.6 (-2.6, -0.7) | -1.7 (-3.3, -0.2) | -1.1 (-2.5, 0.2) | 0.104 |
| Daytime ambulatory SBP | | | | | | | |
| Baseline | 130.0 (12.5) | – | 129.8 (11.5) | – | – | – | – |
| 3 months | 128.5 (11.8) | -1.5 (-3.4, 0.5) | 124.9 (10.7) | -4.9 (-6.6, -3.1) | -3.4 (-7.0, 0.2) | -3.7 (-6.8, -0.7) | 0.018 |
| 9 months | 129.6 (12.3) | -0.4 (-2.4, 1.6) | 126.9 (10.8) | -2.9 (-4.8, -1.1) | -2.5 (-5.5, 0.5) | -2.7 (-5.2, -0.2) | 0.035 |
| Daytime ambulatory DBP | | | | | | | |
| Baseline | 81.7 (7.0) | – | 82.8 (6.8) | – | – | – | – |
| 3 months | 82.4 (7.7) | 0.8 (-0.4, 1.9) | 80.7 (7.5) | -2.0 (-3.1, -1.0) | -2.8 (-4.8, -0.9) | -2.2 (-3.9, -0.5) | 0.013 |
| 9 months | 82.1 (7.1) | 0.4 (-0.6, 1.5) | 81.3 (7.1) | -1.5 (-2.6, -0.5) | -1.9 (-3.7, -0.2) | -1.4 (-2.8, 0.1) | 0.071 |
| Nighttime ambulatory SBP | | | | | | | |
| Baseline | 126.1 (14.4) | – | 126.7 (12.2) | – | – | – | – |
| 3 months | 126.1 (13.1) | 0.0 (-2.2, 2.2) | 123.0 (12.3) | -3.7 (-5.8, -1.6) | -3.7 (-7.4, 0.0) | -3.4 (-6.5, -0.3) | 0.032 |
| 9 months | 126.5 (13.8) | 0.4 (-1.7, 2.5) | 123.6 (12.4) | -3.1 (-5.1, -1.1) | -3.5 (-6.8, -0.3) | -3.2 (-5.9, -0.4) | 0.023 |
| Nighttime ambulatory DBP | | | | | | | |
| Baseline | 79.8 (8.5) | – | 80.7 (7.8) | – | – | – | – |
| 3 months | 80.8 (8.1) | 1.0 (-0.3, 2.3) | 79.4 (7.5) | -1.3 (-2.6, -0.0) | -2.3 (-4.6, -0.1) | -1.7 (-3.6, 0.2) | 0.076 |
| 9 months | 80.4 (7.6) | 0.6 (-0.6, 1.8) | 79.4 (7.7) | -1.3 (-2.5, -0.1) | -1.9 (-3.8, -0.0) | -1.5 (-3.1, 0.1) | 0.065 |
| Office SBP |  |  |  |  |  |  |  |
| Baseline | 130.0 (13.7) | – | 133.3 (13.2) | – | – | – | – |
| 3 months | 126.9 (13.6) | -3.1 (-4.8, -1.4) | 126.7 (12.2) | -6.6 (-8.5, -4.8) | -3.6 (-6.3, -0.9) | -1.9 (-4.3, 0.5) | 0.112 |
| 9 months | 125.4 (13.8) | -4.6 (-6.4, -2.8) | 127.2 (12.8) | -6.1 (-8.0, -4.1) | -1.5 (-4.3, 1.3) | 0.3 (-2.2, 2.8) | 0.811 |
| Office DBP |  |  |  |  |  |  |  |
| Baseline | 82.2 (10.5) | – | 84.7 (10.1) | – | – | – | – |
| 3 months | 80.0 (9.8) | -2.2 (-3.5, -1.0) | 79.3 (9.8) | -5.4 (-6.7, -4.2) | -3.2 (-5.1, -1.3) | -2.0 (-3.7, -0.4) | 0.018 |
| 9 months | 78.6 (9.4) | -3.6 (-4.9, -2.3) | 79.5 (9.4) | -5.2 (-6.5, -4.0) | -1.6 (-3.6, 0.3) | -0.3 (-2.0, 1.4) | 0.721 |
| **Anthropometric** | | | | | | | |
| BMI, kg/m^2^ | | | | | | | |
| Baseline | 26.3 (3.9) | – | 26.1 (3.8) | – | – | – | – |
| 3 months | 26.1 (3.9) | -0.1 (-0.4, 0.1) | 25.8 (3.6) | -0.3 (-0.6, 0.0) | -0.2 (-0.6, 0.3) | -0.1 (-0.6, 0.3) | 0.582 |
| 9 months | 26.2 (4.0) | -0.1 (-0.4, 0.2) | 25.9 (3.6) | -0.3 (-0.6, 0.0) | -0.2 (-0.7, 0.3) | -0.2 (-0.6, 0.3) | 0.472 |
| Fat mass, % | | | | | | | |
| Baseline | 28.6 (7.0) | – | 28.2 (7.0) | – | – | – | – |
| 3 months | 28.4 (6.9) | -0.2 (-0.6, 0.3) | 27.7 (7.0) | -0.5 (-1.0, 0.0) | -0.3 (-1.2, 0.5) | -0.2 (-1.0, 0.6) | 0.572 |
| 9 months | 28.3 (6.8) | -0.3 (-0.7, 0.2) | 27.4 (7.0) | -0.8 (-1.3, -0.2) | -0.5 (-1.3, 0.3) | -0.4 (-1.2, 0.4) | 0.327 |
| **Cardiometabolic Parameters** | | | | | | | |
| Fasting glucose, mmol/L | | | | | | | |
| Baseline | 5.9 (0.9) | – | 6.0 (1.6) | – | – | – | – |
| 3 months | 5.9 (1.3) | 0.1 (-0.1, 0.2) | 6.1 (1.5) | 0.0 (-0.1, 0.1) | -0.1 (-0.3, 0.2) | -0.0 (-0.2, 0.2) | 0.673 |
| 9 months | 6.0 (1.0) | 0.1 (-0.0, 0.2) | 6.1 (2.1) | 0.1 (-0.1, 0.3) | -0.0 (-0.2, 0.2) | -0.0 (-0.2, 0.2) | 0.957 |
| Total cholesterol, mmol/L | | | | | | | |
| Baseline | 4.6 (1.1) | – | 4.8 (1.0) | – | – | – | – |
| 3 months | 4.8 (1.1) | 0.2 (0.1, 0.3) | 4.9 (1.0) | 0.1 (-0.0, 0.2) | -0.1 (-0.3, 0.1) | -0.0 (-0.2, 0.1) | 0.582 |
| 9 months | 4.8 (1.1) | 0.1 (0.0, 0.2) | 4.8 (1.0) | -0.0 (-0.2, 0.1) | -0.2 (-0.4, 0.0) | -0.1 (-0.3, 0.0) | 0.158 |
| LDL-C, mmol/L | | | | | | | |
| Baseline | 2.8 (0.9) | – | 2.9 (0.8) | – | – | – | – |
| 3 months | 2.9 (0.9) | 0.1 (0.0, 0.2) | 3.0 (0.8) | 0.1 (-0.0, 0.2) | -0.0 (-0.2, 0.1) | 0.0 (-0.1, 0.1) | 0.907 |
| 9 months | 2.9 (0.9) | 0.2 (0.1, 0.3) | 2.9 (0.9) | 0.0 (-0.1, 0.1) | -0.2 (-0.3, -0.0) | -0.1 (-0.3, 0.0) | 0.102 |
| HDL-C, mmol/L | | | | | | | |
| Baseline | 1.3 (0.3) | – | 1.4 (0.4) | – | – | – | – |
| 3 months | 1.4 (0.3) | 0.0 (-0.0, 0.1) | 1.4 (0.4) | 0.0 (-0.0, 0.1) | -0.0 (-0.1, 0.0) | 0.0 (-0.0, 0.1) | 0.913 |
| 9 months | 1.3 (0.3) | -0.0 (-0.0, 0.0) | 1.4 (0.4) | -0.0 (-0.0, 0.0) | 0.0 (-0.0, 0.1) | 0.0 (-0.0, 0.1) | 0.509 |
| Triglycerides, mmol/L | | | | | | | |
| Baseline | 1.6 (1.0) | – | 1.6 (1.1) | – | – | – | – |
| 3 months | 1.6 (1.1) | 0.0 (-0.1, 0.2) | 1.5 (0.9) | -0.1 (-0.2, 0.0) | -0.2 (-0.4, 0.1) | -0.2 (-0.4, 0.0) | 0.062 |
| 9 months | 1.7 (1.4) | 0.1 (-0.1, 0.3) | 1.7 (1.6) | 0.1 (-0.1, 0.3) | -0.0 (-0.3, 0.3) | -0.0 (-0.3, 0.2) | 0.746 |
| **Lifestyle** | | | | | | | |
| DASH score | | | | | | | |
| Baseline | 5.9 (1.8) | – | 5.7 (1.8) | – | – | – | – |
| 3 months | 6.4 (1.6) | 0.5 (0.3, 0.7) | 6.8 (1.7) | 1.1 (0.8, 1.3) | 0.6 (0.3, 0.9) | 0.6 (0.3, 0.9) | <0.001 |
| 9 months | 6.5 (1.6) | 0.6 (0.4, 0.8) | 6.8 (1.6) | 1.0 (0.8, 1.3) | 0.5 (0.2, 0.9) | 0.4 (0.2, 0.7) | 0.003 |
| MVPA, MET-min/ week | | | | | | | |
| Baseline | 499.4 (1141.6) | – | 629.8 (1139.0) | – | – | – | – |
| 3 months | 521.7 (1318.0) | 22.3  (-199.9, 244.5) | 860.1 (1136.7) | 230.3  (39.3, 421.3) | 208.0  (-104.8, 520.8) | 324.3  (65.9, 582.8) | 0.014 |
| 9 months | 568.7 (1269.2) | 69.3  (-135.9, 274.4) | 790.6 (1340.1) | 160.8  (-27.6, 349.1) | 91.5  (-212.5, 395.5) | 180.5  (-92.2, 453.2) | 0.194 |
| Walk, MET-min/ week | | | | | | | |
| Baseline | 979.8 (944.1) | – | 991.3  (924.3) | – | – | – | – |
| 3 months | 1012.2 (984.9) | 32.4  (-124.7, 189.6) | 1043.3 (962.3) | 52.0  (-114.8, 218.8) | 19.6  (-229.3, 268.4) | 10.3  (-199.2, 219.8) | 0.923 |
| 9 months | 892.2 (834.0) | -87.6  (-234.4, 59.1) | 970.4  (890.7) | -20.9  (-162.3, 120.5) | 66.7  (-152.4, 285.8) | 78.8  (-98.5, 256.2) | 0.382 |
| Daily step counts, steps | | | | | | | |
| 0–3 months | 6956.3  (3574.2) | – | 7107.9  (3156.8) | – | 151.7  (-905.9, 602.6) | 129.5  (-626, 885) | 0.736 |
| 4–9 months | 5899.9  (3569) | – | 5953.3  (3109.9) | – | 53.4  (-791.7, 685) | 40.1  (-701.8, 782) | 0.915 |

* General linear models with adjustment for baseline outcome, blood pressure category, study center, age, sex, education level, per capita income monthly, smoking status.

Daily step counts were summarized for months 0–3 and months 4–9 as absolute levels. EUC indicates enhanced usual care; Tele-LM, telehealth-based lifestyle modification program; VO_2peak_, peak oxygen uptake; HRmax, maximal heart rate; RER, respiratory exchange ratio; SBP, systolic blood pressure; DBP, diastolic blood pressure; BMI, body mass index; LDL-C, low-density lipoprotein cholesterol; HDL-C, high-density lipoprotein cholesterol; DASH, dietary approaches to stop hypertension; MVPA, moderate to vigorous physical activity; MET, metabolic equivalent task.

### **Table S3. Subgroup Analyses According to Randomization Strata (Blood Pressure Category) for Between-group Differences in Changes for Primary and Secondary Outcomes at 3 Months**

| **Outcomes** | **High-normal blood pressure** | | | | **High blood pressure** | | | | ***P* for interaction**^†^ |
| --- | --- | --- | --- | --- | --- | --- | --- | --- | --- |
|  | **EUC**  **(N=64)** | **Tele-LM**  **(N=65)** | **Difference (95% CI)^*^** | ***P*-value** | **EUC**  **(N=148)** | **Tele-LM**  **(N=147)** | **Difference (95% CI)^*^** | ***P*-value** |  |
| **VO_2peak_, mL/kg/min** | -0.7  (-2.0, 0.7) | 1.5  (0.1, 2.9) | 2.1  (0.3, 4.0) | 0.026 | 0.0  (-0.7, 0.7) | 1.2  (0.4, 2.0) | 1.3  (0.2, 2.3) | 0.015 | 0.393 |
| **Home SBP, mmHg** | -3.6  (-6.6, -0.5) | -7.0  (-9.7, -4.4) | -2.8  (-6.2, 0.5) | 0.094 | -1.7  (-3.8, 0.4) | -5.1  (-7.2, -3.0) | -3.3  (-5.6, -1.0) | 0.006 | 0.804 |
| **Home DBP, mmHg** | -0.6  (-2.5, 1.2) | -3.1  (-5.0, -1.3) | -1.9  (-4.3, 0.5) | 0.112 | 1.1  (-0.2, 2.4) | -1.8  (-3.1, -0.5) | -2.8  (-4.3, -1.2) | <0.001 | 0.506 |
| **24-hour ambulatory SBP, mmHg** | -2.3  (-6.2, 1.5) | -8.0  (-12.1, -3.9) | -4.3  (-9.0, 0.4) | 0.073 | -1.1  (-4.0, 1.8) | -4.0  (-6.3, -1.6) | -2.7  (-6.0, 0.6) | 0.106 | 0.553 |
| **24-hour ambulatory DBP, mmHg** | -0.1  (-2.5, 2.2) | -3.2  (-5.6, -0.8) | -2.1  (-5.0, 0.8) | 0.157 | 0.7  (-0.9, 2.4) | -1.8  (-3.3, -0.3) | -1.8  (-3.8, 0.2) | 0.070 | 0.880 |
| **Office SBP, mmHg** | -5.2  (-8.1, -2.3) | -7.5  (-11.3, -3.7) | 0.2  (-4.1, 4.5) | 0.917 | -2.1  (-4.4, 0.1) | -6.2  (-8.6, -3.9) | -2.7  (-5.5, 0.0) | 0.053 | 0.228 |
| **Office DBP, mmHg** | -3.5  (-5.9, -1.0) | -4.8  (-7.3, -2.3) | 0.7  (-2.4, 3.9) | 0.641 | -1.7  (-3.3, -0.1) | -5.7  (-7.3, -4.1) | -3.2  (-5.2, -1.1) | 0.002 | 0.043 |

Values are mean changes (95% CI) or between-group differences in changes (95% CI). Positive differences for VO_2peak_ and negative differences for blood pressure are in favor of the Tele-LM.

* General linear models with adjustment for baseline values of the dependent variable.

† Tests for interaction between intervention assignment and blood pressure category.

EUC indicates enhanced usual care; Tele-LM, telehealth-based lifestyle modification program; VO_2peak_, peak oxygen uptake; SBP, systolic blood pressure; DBP, diastolic blood pressure.

### **Table S4. Subgroup Analyses for Primary and Secondary Outcomes in EUC and Tele-LM at 3 Months.**

| **Subgroup** | **N_EUC** | **N_Tele-LM** | **Difference**  **(95% CI)^*^** | ***P*-value** | ***P* for interaction**^†^ |
| --- | --- | --- | --- | --- | --- |
| VO_2peak_, mL/kg/min | | | | | |
| Age, years |  |  |  |  | 0.863 |
| <50 | 98 | 103 | 1.6 (0.1, 3.0) | 0.036 |  |
| ≥50 | 114 | 109 | 1.4 (0.3, 2.5) | 0.012 |  |
| Sex |  |  |  |  | 0.572 |
| Male | 134 | 142 | 1.7 (0.5, 2.9) | 0.007 |  |
| Female | 78 | 70 | 1.1 (-0.2, 2.4) | 0.099 |  |
| Center |  |  |  |  | 0.791 |
| Beijing | 185 | 194 | 1.5 (0.6, 2.4) | 0.001 |  |
| Shenzhen | 27 | 18 | 1.7 (-1.5, 4.9) | 0.286 |  |
| Smoking status |  |  |  |  | 0.540 |
| Current | 38 | 44 | 1.0 (-1.1, 3.1) | 0.348 |  |
| Former | 25 | 19 | 2.7 (-0.5, 6.0) | 0.091 |  |
| Never | 149 | 149 | 1.5 (0.4, 2.6) | 0.007 |  |
| Home SBP, mmHg | | | | | |
| Age, years |  |  |  |  | 0.474 |
| <50 | 98 | 103 | -2.4 (-5.3, 0.5) | 0.109 |  |
| ≥50 | 114 | 109 | -3.8 (-6.4, -1.2) | 0.005 |  |
| Sex |  |  |  |  | 0.317 |
| Male | 134 | 142 | -2.5 (-5.0, 0.1) | 0.056 |  |
| Female | 78 | 70 | -4.5 (-7.6, -1.5) | 0.004 |  |
| Center |  |  |  |  | 0.866 |
| Beijing | 185 | 194 | -3.3 (-5.3, -1.2) | 0.002 |  |
| Shenzhen | 27 | 18 | -3.1 (-8.6, 2.3) | 0.249 |  |
| Smoking status |  |  |  |  | 0.951 |
| Current | 38 | 44 | -3.1 (-8.5, 2.3) | 0.254 |  |
| Former | 25 | 19 | -1.9 (-9.5, 5.6) | 0.597 |  |
| Never | 149 | 149 | -3.4 (-5.4, -1.4) | 0.001 |  |
| Home DBP, mmHg | | | | | |
| Age, years |  |  |  |  | 0.354 |
| <50 | 98 | 103 | -1.7 (-3.8, 0.3) | 0.098 |  |
| ≥50 | 114 | 109 | -3.1 (-4.8, -1.4) | <0.001 |  |
| Sex |  |  |  |  | 0.481 |
| Male | 134 | 142 | -2.2 (-3.9, -0.5) | 0.011 |  |
| Female | 78 | 70 | -3.2 (-5.2, -1.1) | 0.003 |  |
| Center |  |  |  |  | 0.939 |
| Beijing | 185 | 194 | -2.5 (-3.9, -1.1) | <0.001 |  |
| Shenzhen | 27 | 18 | -2.3 (-5.9, 1.4) | 0.209 |  |
| Smoking status |  |  |  |  | 0.701 |
| Current | 38 | 44 | -2.7 (-6.1, 0.7) | 0.114 |  |
| Former | 25 | 19 | -0.7 (-5.9, 4.4) | 0.773 |  |
| Never | 149 | 149 | -2.7 (-4.1, -1.4) | <0.001 |  |
| 24-hour ambulatory SBP, mmHg | | | | | |
| Age, years |  |  |  |  | 0.430 |
| <50 | 98 | 103 | -4.2 (-8.2, -0.3) | 0.036 |  |
| ≥50 | 114 | 109 | -2.2 (-5.9, 1.6) | 0.253 |  |
| Sex |  |  |  |  | 0.631 |
| Male | 134 | 142 | -2.7 (-6.1, 0.6) | 0.109 |  |
| Female | 78 | 70 | -4.0 (-8.5, 0.4) | 0.075 |  |
| Center |  |  |  |  | 0.788 |
| Beijing | 185 | 194 | -3.4 (-6.3, -0.5) | 0.022 |  |
| Shenzhen | 27 | 18 | -2.6 (-10.0, 4.7) | 0.463 |  |
| Smoking status |  |  |  |  | 0.979 |
| Current | 38 | 44 | -3.6 (-10.8, 3.6) | 0.321 |  |
| Former | 25 | 19 | -4.0 (-13.1, 5.0) | 0.366 |  |
| Never | 149 | 149 | -3.0 (-6.1, 0.1) | 0.062 |  |
| 24-hour ambulatory DBP, mmHg | | | | | |
| Age, years |  |  |  |  | 0.418 |
| <50 | 98 | 103 | -2.5 (-4.9, -0.1) | 0.045 |  |
| ≥50 | 114 | 109 | -1.3 (-3.5, 0.8) | 0.231 |  |
| Sex |  |  |  |  | 0.748 |
| Male | 134 | 142 | -1.8 (-3.9, 0.3) | 0.098 |  |
| Female | 78 | 70 | -2.3 (-4.8, 0.3) | 0.078 |  |
| Center |  |  |  |  | 0.810 |
| Beijing | 185 | 194 | -2.0 (-3.7, -0.2) | 0.027 |  |
| Shenzhen | 27 | 18 | -1.1 (-5.6, 3.3) | 0.607 |  |
| Smoking status |  |  |  |  | 0.981 |
| Current | 38 | 44 | -1.7 (-5.7, 2.3) | 0.393 |  |
| Former | 25 | 19 | -2.8 (-9.3, 3.6) | 0.371 |  |
| Never | 149 | 149 | -1.8 (-3.6, 0.0) | 0.051 |  |
| Office SBP, mmHg | | | | | |
| Age, years |  |  |  |  | 0.935 |
| <50 | 98 | 103 | -1.3 (-4.6, 2.0) | 0.426 |  |
| ≥50 | 114 | 109 | -2.0 (-5.4, 1.3) | 0.229 |  |
| Sex |  |  |  |  | 0.507 |
| Male | 134 | 142 | -1.1 (-3.9, 1.8) | 0.463 |  |
| Female | 78 | 70 | -3.5 (-7.6, 0.5) | 0.085 |  |
| Center |  |  |  |  | 0.690 |
| Beijing | 185 | 194 | -1.7 (-4.2, 0.9) | 0.193 |  |
| Shenzhen | 27 | 18 | -3.6 (-8.8, 1.7) | 0.175 |  |
| Smoking status |  |  |  |  | 0.559 |
| Current | 38 | 44 | 0.4 (-5.0, 5.8) | 0.877 |  |
| Former | 25 | 19 | -3.2 (-10.9, 4.5) | 0.399 |  |
| Never | 149 | 149 | -2.4 (-5.2, 0.5) | 0.100 |  |
| Office DBP, mmHg | | | | | |
| Age, years |  |  |  |  | 0.758 |
| <50 | 98 | 103 | -1.7 (-4.4, 1.0) | 0.214 |  |
| ≥50 | 114 | 109 | -2.3 (-4.4, -0.2) | 0.034 |  |
| Sex |  |  |  |  | 0.238 |
| Male | 134 | 142 | -1.3 (-3.5, 0.9) | 0.242 |  |
| Female | 78 | 70 | -3.5 (-6.1, -0.8) | 0.010 |  |
| Center |  |  |  |  | 0.371 |
| Beijing | 185 | 194 | -1.7 (-3.6, 0.1) | 0.060 |  |
| Shenzhen | 27 | 18 | -4.5 (-9.6, 0.6) | 0.083 |  |
| Smoking status |  |  |  |  | 0.806 |
| Current | 38 | 44 | -1.1 (-5.5, 3.3) | 0.623 |  |
| Former | 25 | 19 | -3.1 (-9.0, 2.8) | 0.289 |  |
| Never | 149 | 149 | -2.1 (-4.0, -0.2) | 0.031 |  |

Positive differences for VO_2peak_ and negative differences for blood pressure are in favor of the Tele-LM.

* General linear models with adjustment for baseline values of the dependent variable.

† Tests for interaction between intervention assignment and each subgroup.

EUC indicates enhanced usual care; Tele-LM, telehealth-based lifestyle modification program; VO_2peak_, peak oxygen uptake; SBP, systolic blood pressure; DBP, diastolic blood pressure.

### **Table S5. Safety Outcomes and Adverse Events by Participants During the Trial Period**

| **Any adverse event** | **EUC (N=212)** | **Tele-LM (N=212)** |
| --- | --- | --- |
| Cardiovascular disease | 1 | 1 |
| Poorly controlled hypertension | 2 | 0 |
| Musculoskeletal pain | 0 | 2 |
| Leg injury | 0 | 1 |
| Anterior cruciate ligament tear | 1 | 0 |
| Cough | 1 | 0 |
| Right wrist fracture | 1 | 0 |
| Left arm fracture | 0 | 1 |
| Left foot fracture | 0 | 1 |
| Bruise | 0 | 1 |
| Thyroid nodule | 0 | 1 |
| Sudden deafness | 0 | 1 |
| Fasciitis | 1 | 0 |
| Rib fracture | 0 | 1 |
| Pneumonia | 1 | 0 |
| Periostitis | 1 | 0 |

EUC indicates enhanced usual care; Tele-LM, telehealth-based lifestyle modification program.

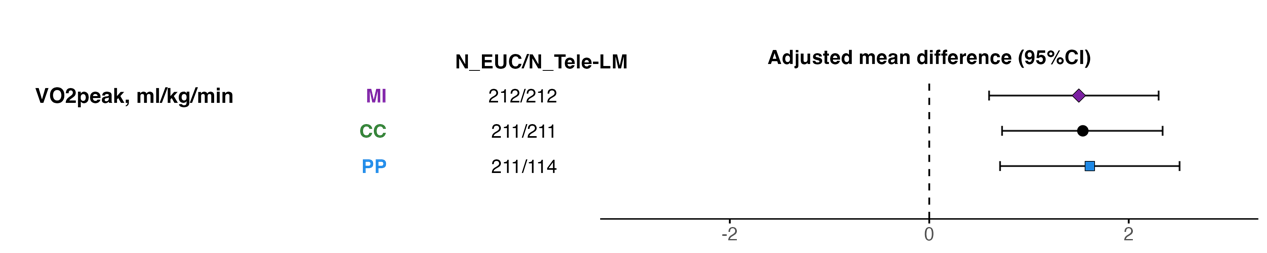

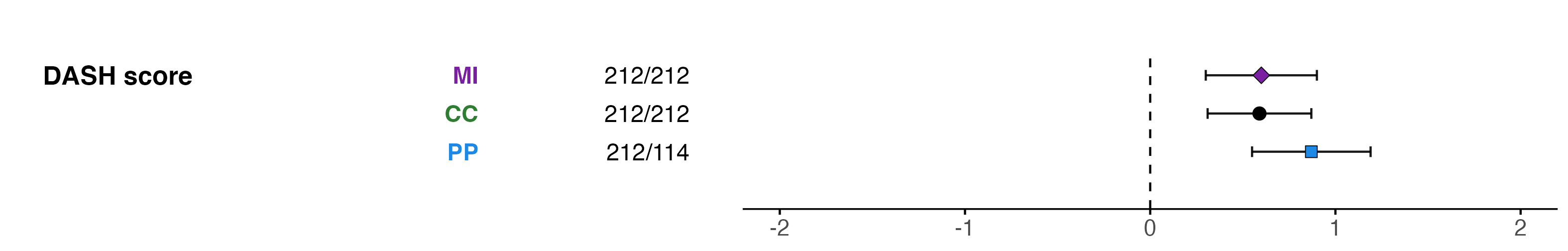

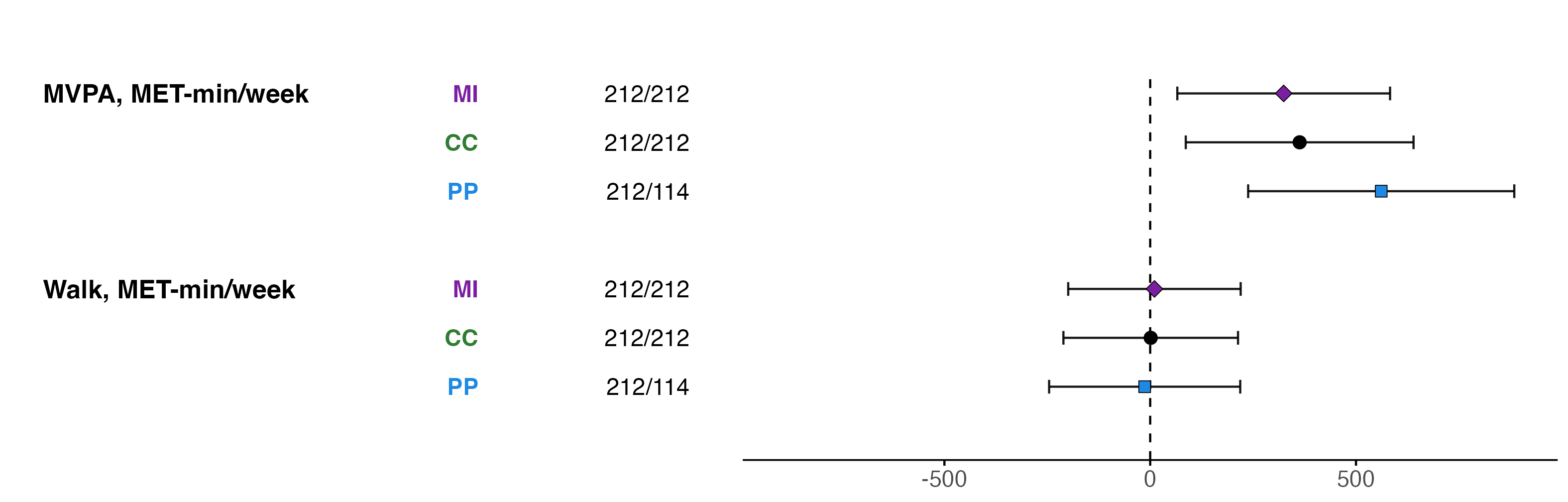

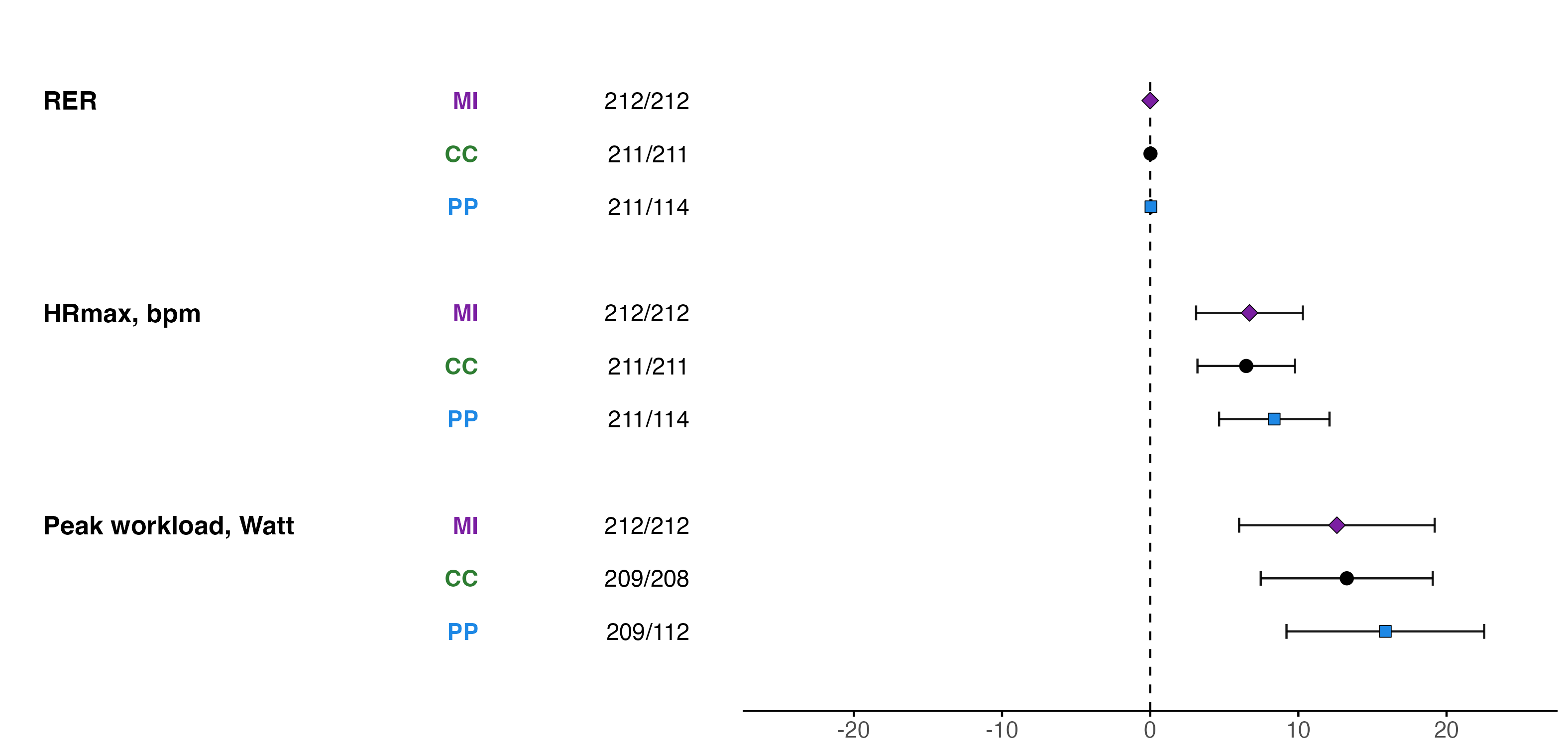

### **Figure S1. Sensitivity Analyses of Cardiorespiratory Fitness and Lifestyle Outcomes for MI, CC and PP at 3 months**

Positive differences are in favor of the Tele-LM. The MI analysis followed the intention-to-treat principle with multiple imputation for missing data. The CC analysis included all participants with available data at baseline and 3 months for the respective endpoint. The PP analysis included all participants randomized to the EUC and those participants randomized to the Tele-LM who performed at least 50% of prescribed training duration.

MI indicates multiple imputation; CC, complete-case; PP, per-protocol; EUC, enhanced usual care; Tele-LM, telehealth-based lifestyle modification program; VO_2peak_, peak oxygen uptake; RER, respiratory exchange ratio; HRmax, maximal heart rate; MVPA, moderate to vigorous physical activity; MET, metabolic equivalent task; DASH, dietary approaches to stop hypertension.

### **Figure S2. Sensitivity Analyses of Blood Pressure Outcomes for MI, CC and PP at 3 months**

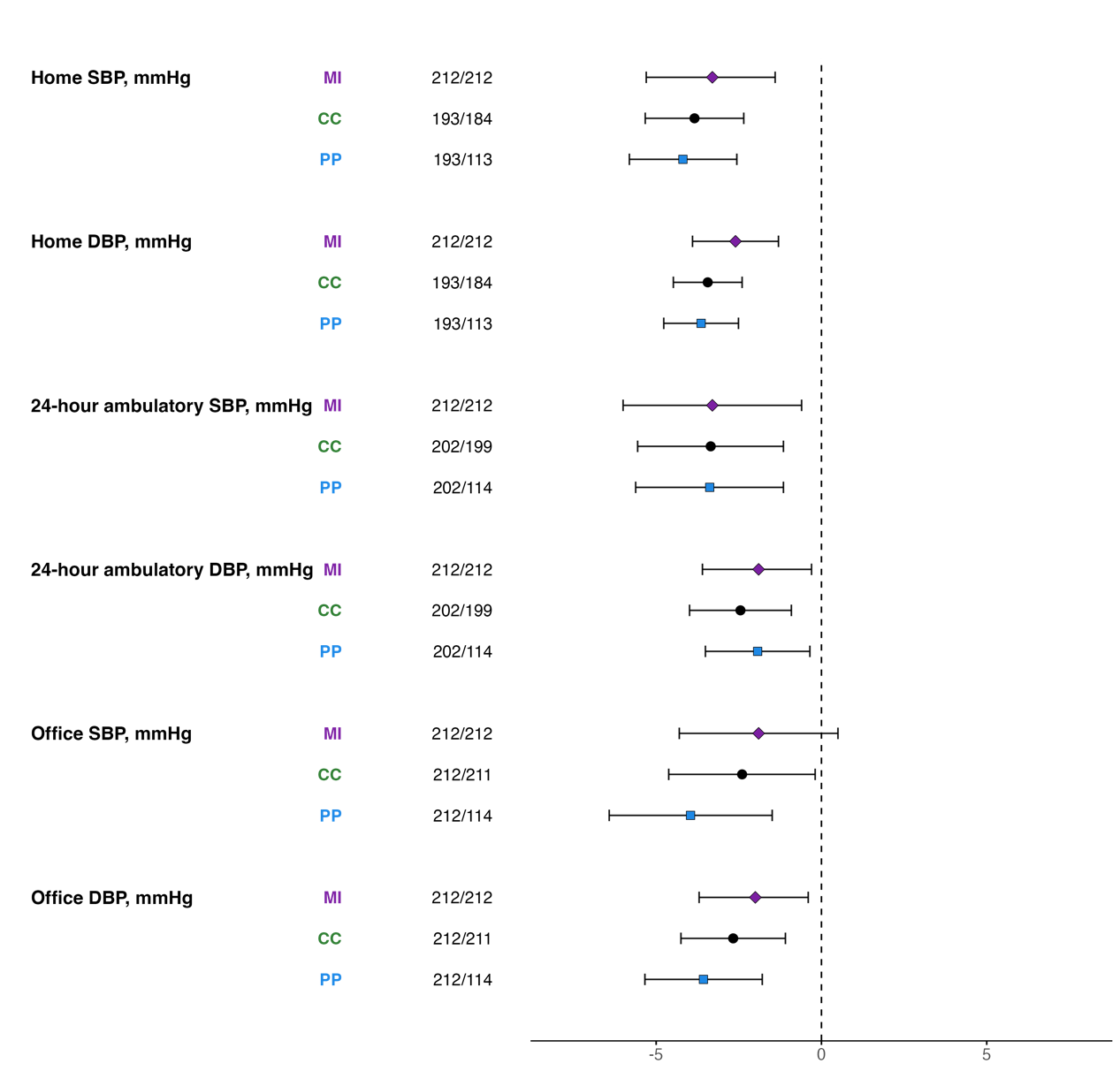

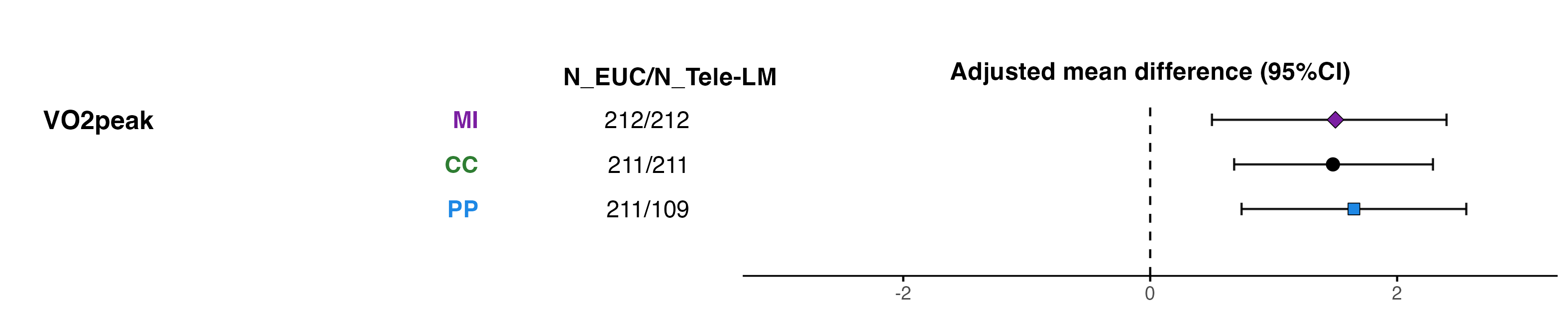

Negative differences are in favor of the Tele-LM. The MI analysis followed the intention-to-treat principle with multiple imputation for missing data. The CC analysis included all participants with available data at baseline and 3 months for the respective endpoint. The PP analysis included all participants randomized to the EUC and those participants randomized to the Tele-LM who performed at least 50% of prescribed training duration.

MI indicates multiple imputation; CC, complete-case; PP, per-protocol; EUC, enhanced usual care; Tele-LM, telehealth-based lifestyle modification program; SBP, systolic blood pressure; DBP, diastolic blood pressure.

### **Figure S3. Sensitivity Analyses of Anthropometric and Cardiometabolic Parameters Outcomes for MI, CC and PP at 3 months**

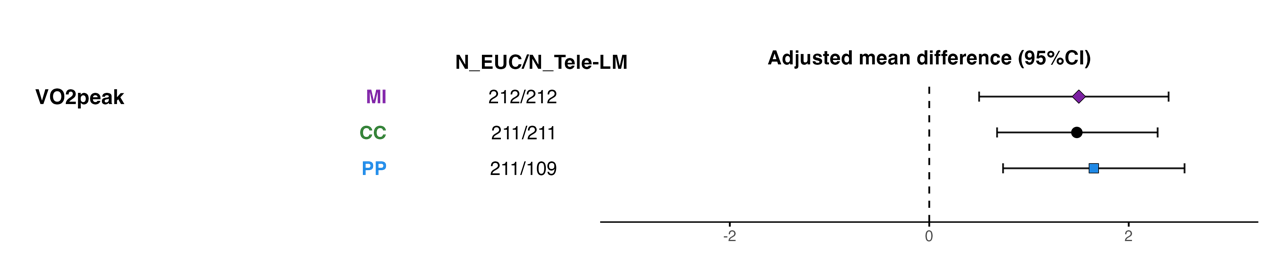

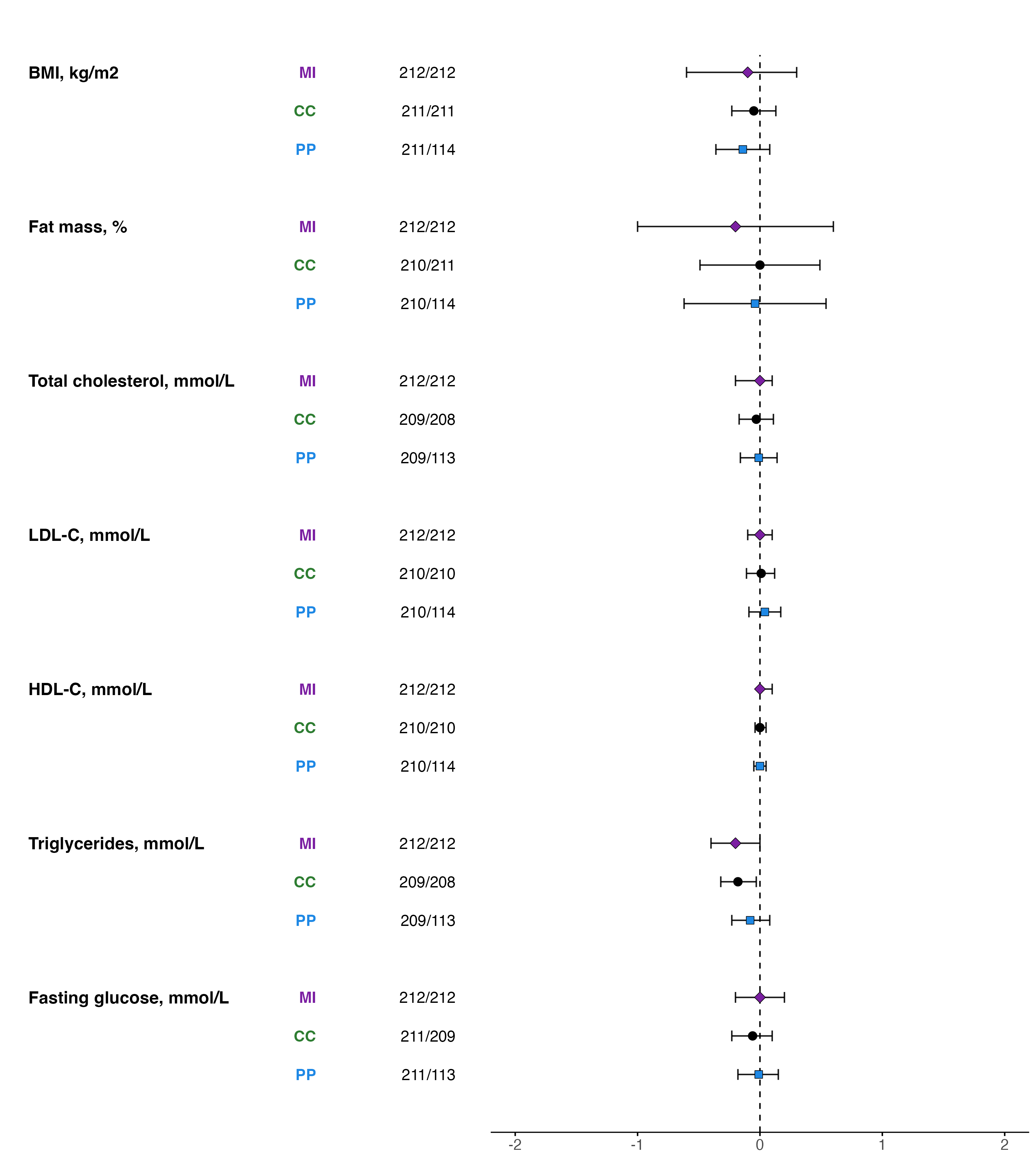

Negative differences are in favor of the Tele-LM except for HDL-C, where positive differences indicate benefit. The MI analysis followed the intention-to-treat principle with multiple imputation for missing data. The CC analysis included all participants with available data at baseline and 3 months for the respective endpoint. The PP analysis included all participants randomized to the EUC and those participants randomized to the Tele-LM who performed at least 50% of prescribed training duration.

MI indicates multiple imputation; CC, complete-case; PP, per-protocol; EUC, enhanced usual care; Tele-LM, telehealth-based lifestyle modification program; BMI, body mass index; LDL-C, low-density lipoprotein cholesterol; HDL-C, high-density lipoprotein cholesterol.

### **Figure S4. Sensitivity Analyses of Cardiorespiratory Fitness and Lifestyle Outcomes for MI, CC and PP at 9 months**

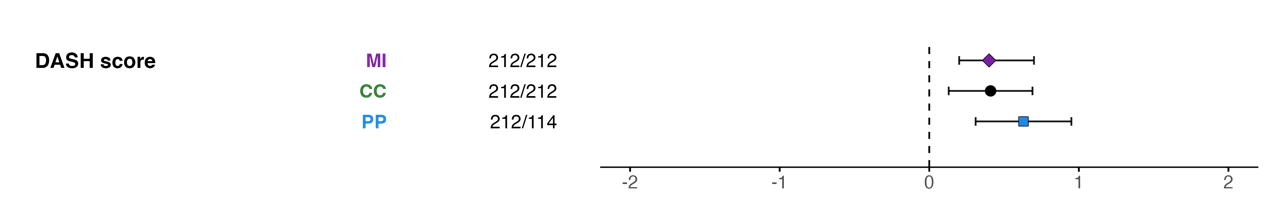

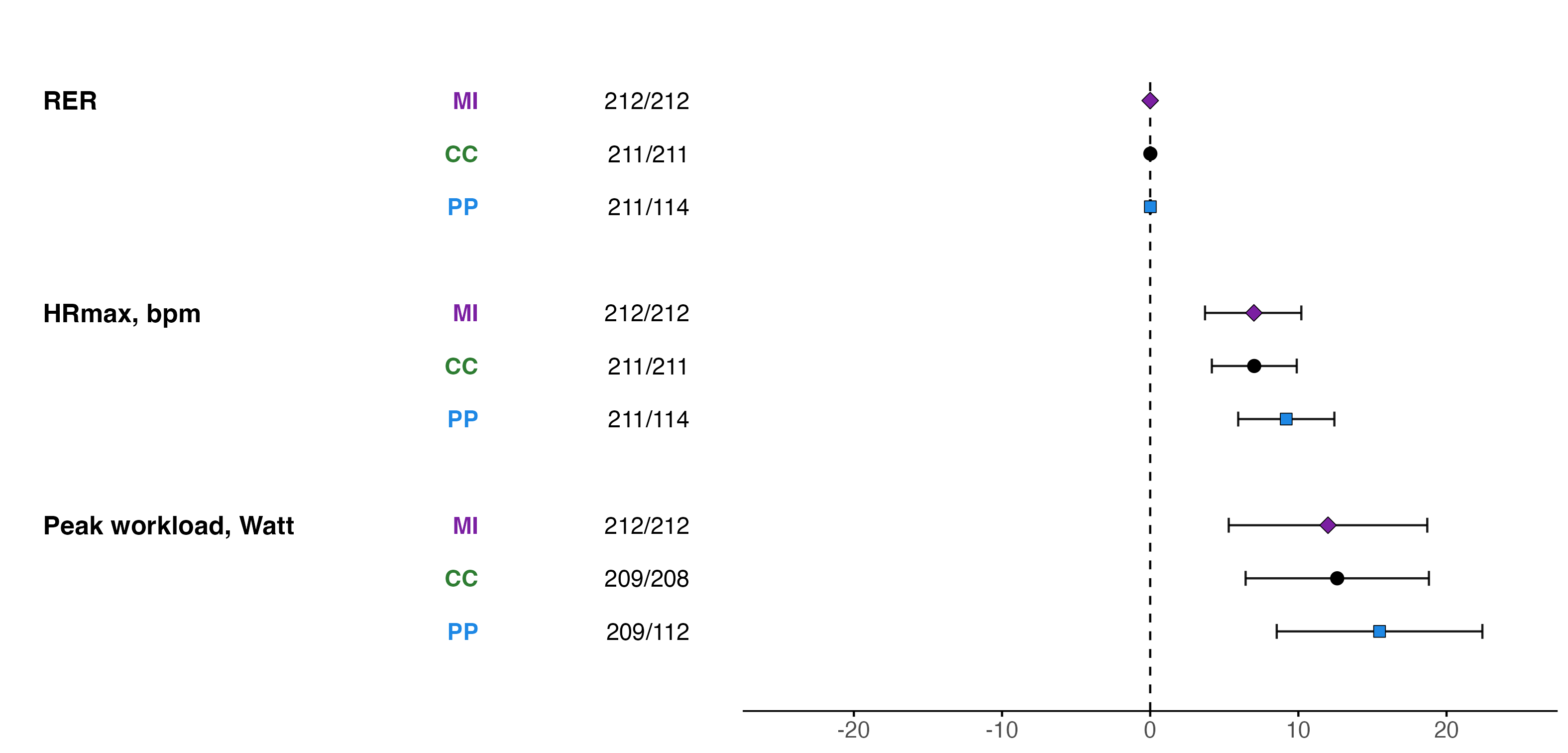

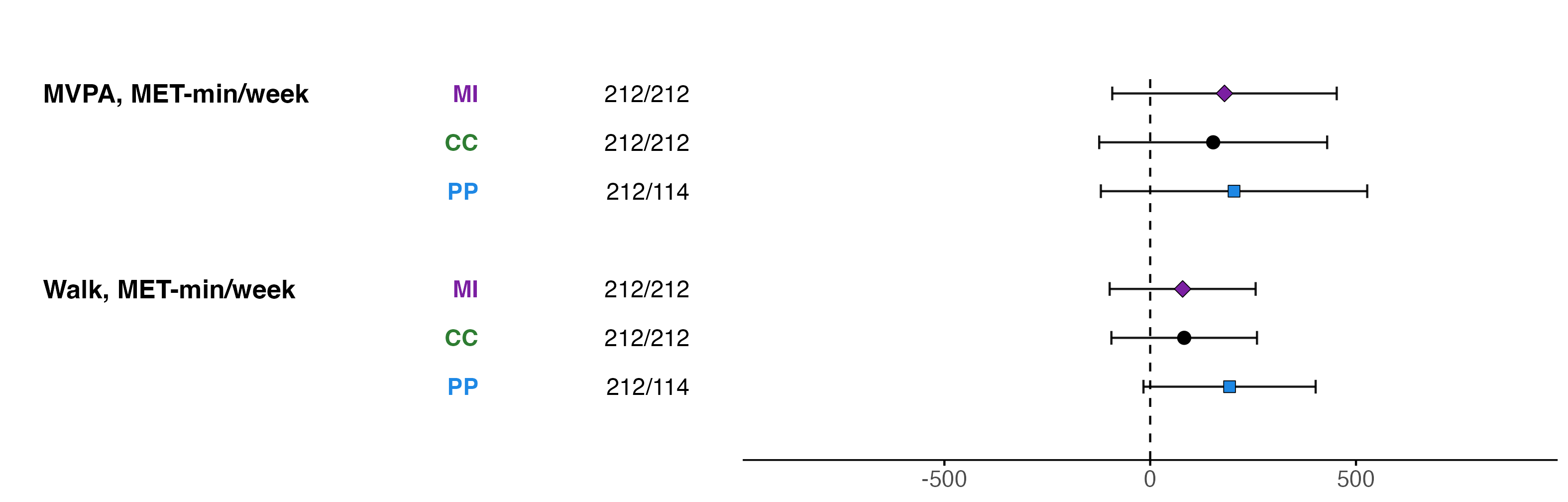

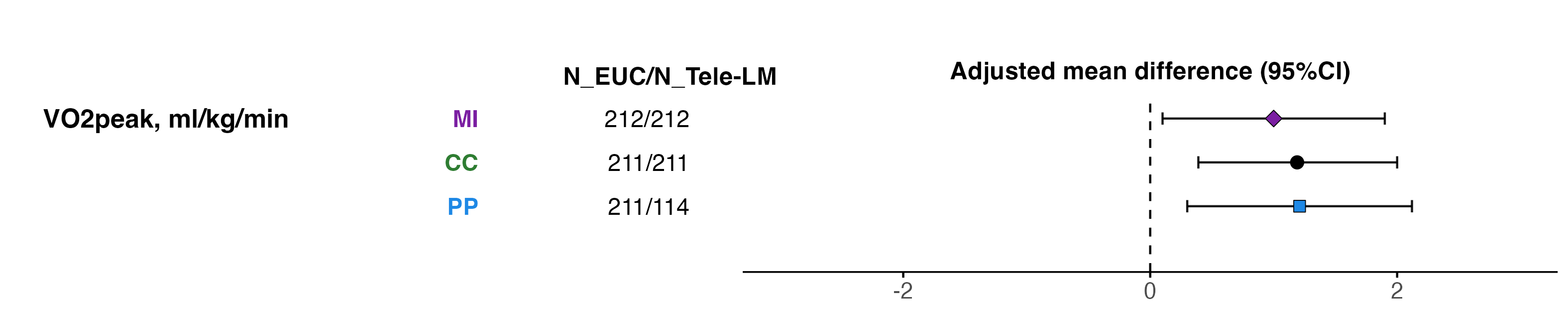

Positive differences are in favor of the Tele-LM. The MI analysis followed the intention-to-treat principle with multiple imputation for missing data. The CC analysis included all participants with available data at baseline and 9 months for the respective endpoint. The PP analysis included all participants randomized to the EUC and those participants randomized to the Tele-LM who performed at least 50% of prescribed training duration.

MI indicates multiple imputation; CC, complete-case; PP, per-protocol; EUC, enhanced usual care; Tele-LM, telehealth-based lifestyle modification program; VO_2peak_, peak oxygen uptake; RER, respiratory exchange ratio; HRmax, maximal heart rate; MVPA, moderate to vigorous physical activity; MET, metabolic equivalent task; DASH, dietary approaches to stop hypertension.

### **Figure S5. Sensitivity Analyses of Blood Pressure Outcomes for MI, CC and PP at 9 months**

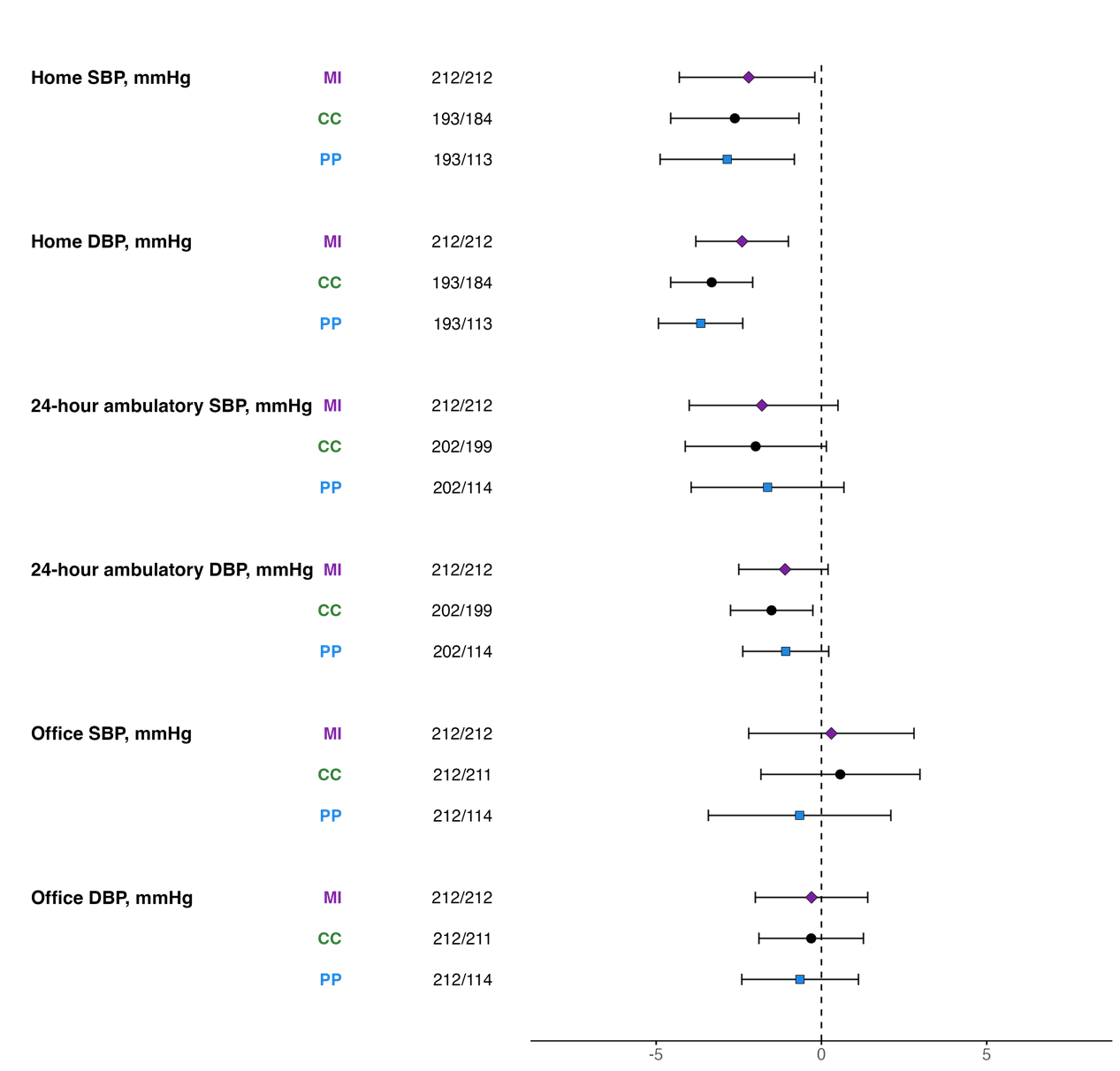

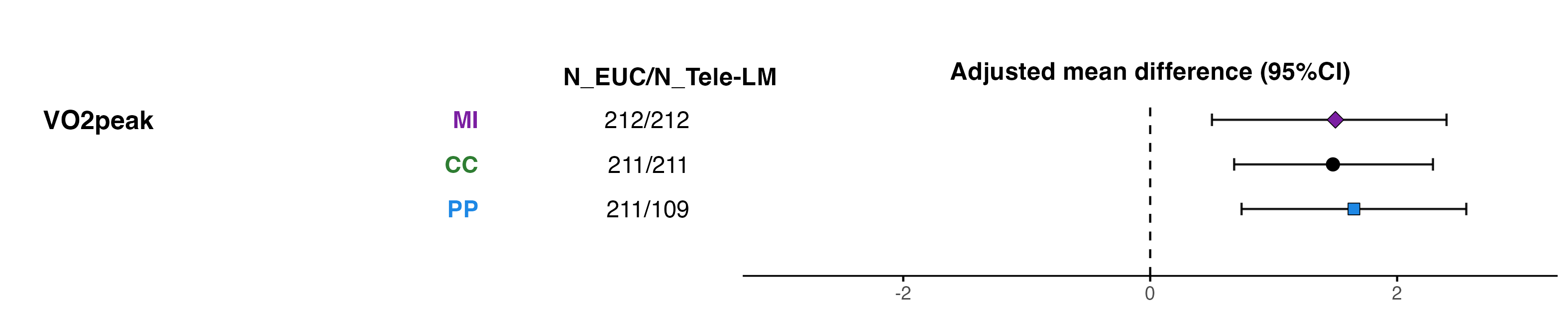

Negative differences are in favor of the Tele-LM. The MI analysis followed the intention-to-treat principle with multiple imputation for missing data. The CC analysis included all participants with available data at baseline and 9 months for the respective endpoint. The PP analysis included all participants randomized to the EUC and those participants randomized to the Tele-LM who performed at least 50% of prescribed training duration.

MI indicates multiple imputation; CC, complete-case; PP, per-protocol; EUC, enhanced usual care; Tele-LM, telehealth-based lifestyle modification program; SBP, systolic blood pressure; DBP, diastolic blood pressure.

### **Figure S6. Sensitivity Analyses of Anthropometric and Cardiometabolic Parameters Outcomes for MI, CC and PP at 9 months**

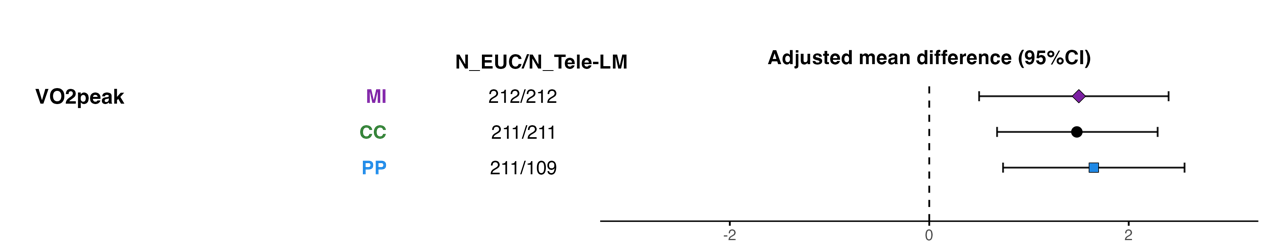

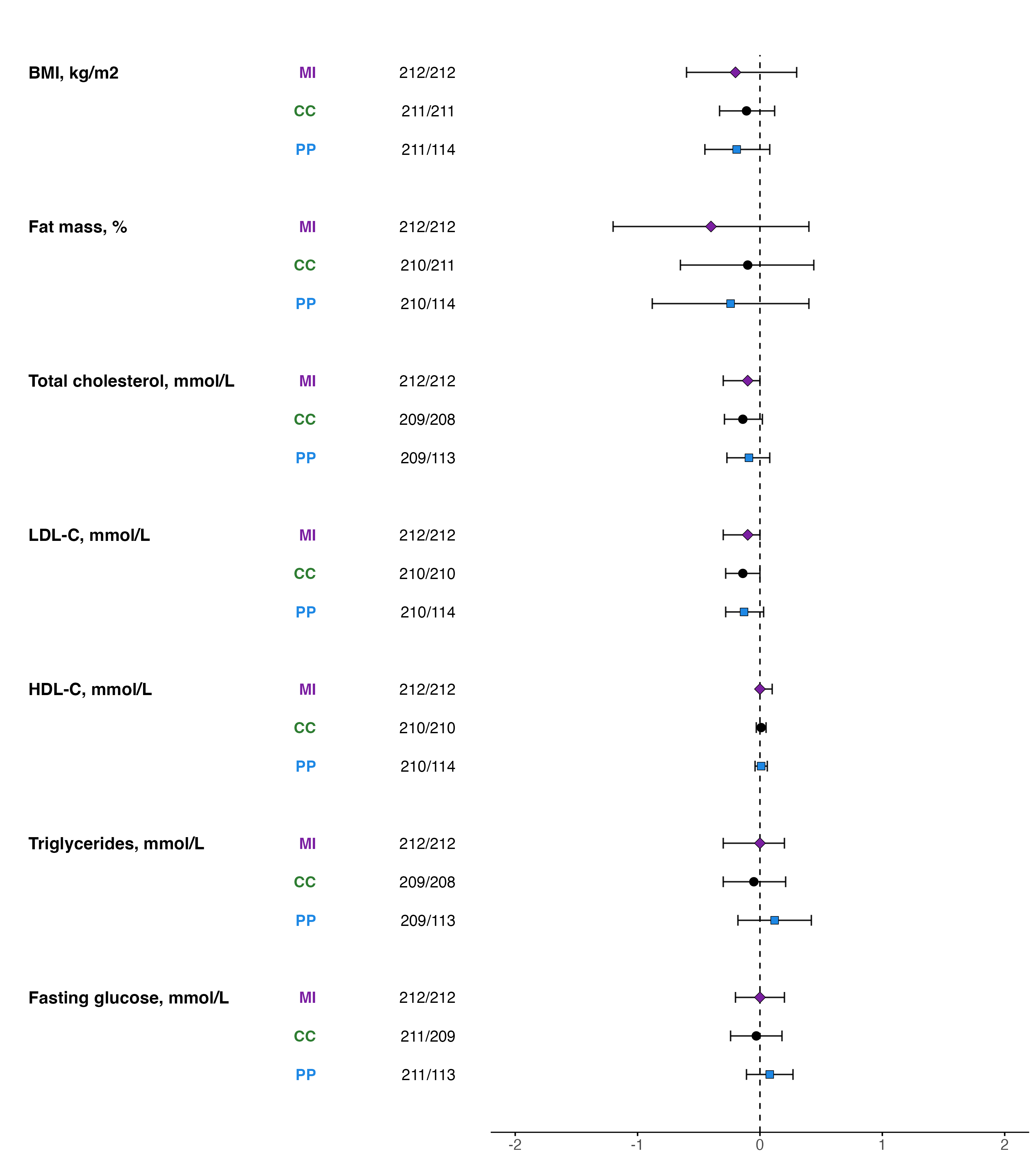

Negative differences are in favor of the Tele-LM except for HDL-C, where positive differences indicate benefit. The MI analysis followed the intention-to-treat principle with multiple imputation for missing data. The CC analysis included all participants with available data at baseline and 9 months for the respective endpoint. The PP analysis included all participants randomized to the EUC and those participants randomized to the Tele-LM who performed at least 50% of prescribed training duration.

MI indicates multiple imputation; CC, complete-case; PP, per-protocol; EUC, enhanced usual care; Tele-LM, telehealth-based lifestyle modification program; BMI, body mass index; LDL-C, low-density lipoprotein cholesterol; HDL-C, high-density lipoprotein cholesterol.
